# Complexity of the neutrophil transcriptome in early and severe rheumatoid arthritis. A role for microRNAs?

**DOI:** 10.1101/2024.12.12.24318900

**Authors:** Michele Fresneda Alarcon, Genna Ali Abdullah, John Alexander Beggs, Isobel Kynoch, Andrew Sellin, Andrew Cross, Sam Haldenby, Philipp Antczak, Eva Caamaño Gutiérrez, Helen Louise Wright

**Author notes:** contributed equally (Michele Fresneda Alarcon, Genna Ali Abdullah).

## Abstract

Neutrophils are innate immune cells that drive the progression of rheumatoid arthritis (RA) through the release of reactive oxygen species (ROS), neutrophil extracellular traps (NETs) and proteases that damage host tissues. Neutrophil activation is regulated, in part, by dynamic changes in gene expression. In this study we have used RNAseq to measure the transcriptomes of neutrophils from people with severe, methotrexate-refractory RA and healthy controls. We identified a dynamic gene expression profile in people with severe RA. This is dominated by a type-I interferon-induced gene expression signature as well as activation of genes regulating neutrophil degranulation, NET production, response to ROS and oxidative stress. Whilst we did not detect significantly elevated levels of interferon-alpha in RA blood sera, we identified increased expression in RA neutrophils of miR-96- 5p and miR-183-5p which regulate activation of the interferon pathway as members of the miR-183C cluster. We also detected significantly elevated NET debris in RA blood sera (p<0.05). Using gene set variation analysis we explored the heterogeneity of neutrophil gene expression in RA and identified subsets of patients with gene expression profiles reflecting enhanced neutrophil degranulation and cytotoxicity, tissue inflammation or activation by interferons. Comparison with published single-cell RNAseq datasets identified RA transcriptomes where neutrophils were polarised by genes relating to early or late cell maturity, with significant genes in each polarised state being regulated by miR-146a- 5p, miR-155-5p, miR-183-5p or miR-96-5p. Overall our study demonstrates the heterogeneity of the RA neutrophil transcriptome and proposes miRNA-driven mechanisms for regulating the activated neutrophil phenotype in RA.

## 1. INTRODUCTION

Rheumatoid arthritis (RA) is a chronic autoimmune disorder characterized by persistent inflammation and the progressive destruction of joint tissues. While the exact aetiology of RA remains unclear, it is well understood that the disease involves a complex interplay of genetic, environmental and immunological factors [1]. Among the key players in the pathogenesis of RA are neutrophils, which are the most abundant type of blood leukocyte in humans and play a crucial role in the innate immune response [2, 3]. Neutrophils are traditionally recognised for their role in the rapid response to infections, being the first immune cells to arrive at sites of inflammation. They can engulf and destroy pathogens through phagocytosis, release antimicrobial proteases and generate extracellular traps (NETs) to immobilize and kill microbes [1]. However, in autoimmune diseases like RA, the dysregulated activation of neutrophils contributes to the pathology of the disease rather than resolving any infection [1, 4].

In RA, neutrophils are excessively activated and accumulate in the synovial fluid and tissue of affected joints [5, 6]. This accumulation not only reflects an increased influx but also decreased apoptosis, leading to prolonged survival of these cells in the joint space [7, 8]. Activated neutrophils in the RA synovium release a range of pro-inflammatory mediators, including cytokines interleukin-1 (IL-1) and tumour necrosis factor-alpha (TNF-α) as well as a number of chemokines, which further amplify the inflammatory response [9, 10]. Additionally, they produce reactive oxygen species (ROS) and proteolytic enzymes including collagenase, contributing to tissue damage and the erosion of cartilage and bone [3, 6, 11–13].

Recent advancements in molecular biology and genomics have shed light on the gene expression profiles of neutrophils in RA, particularly during inflammation. Gene expression profiling studies have shown that neutrophils from people with RA exhibit distinct transcriptional signatures compared to those from healthy individuals [14, 15]. These signatures are characterized by the upregulation of genes involved in inflammatory responses, migration and survival [16]. For instance, genes encoding components of the NADPH oxidase (NOX2) complex, which plays a critical role in the production of ROS, are often upregulated in RA neutrophils. Furthermore, the interaction between neutrophils and other immune cells, such as T cells and macrophages, is also influenced by their gene expression. The expression of surface molecules that modulate immune cell interactions, such as adhesion molecules and antigen-presenting molecules, is altered in RA neutrophils [16]. This can affect the recruitment and activation of other immune cells, contributing to the perpetuation of the inflammatory process [17–19].

Another important aspect of RA neutrophils is their role in forming neutrophil extracellular traps (NETs), which are formed from externalised DNA, histones and neutrophil-derived proteins. In RA, the formation of NETs is thought to contribute to the autoimmune response by exposing intracellular antigens to the immune system [20, 21]. Studies have shown that NETs are abundant in the synovial fluid of RA patients and may be involved in the formation of autoantibodies typical of RA, such as anti-citrullinated protein antibodies (ACPAs) [6, 20, 22]. We have also recently identified a sub-population of DEspR positive, NET-forming neutrophils in RA blood smears [23].

In this study we aim to further delineate the heterogeneity of gene expression profile of RA neutrophils and propose inflammatory factors regulating gene expression and production of pro-inflammatory molecules by neutrophils. We also demonstrate altered expression of microRNAs involved in regulating neutrophil mRNA expression. Understanding the specific pathways and molecular mechanisms underlying the abnormal behaviour of neutrophils in RA could open new avenues for targeted therapies, potentially leading to better management and treatment of this debilitating condition.

## 2. METHODS

### 2.1 Ethics and patients

This study was approved by the University of Liverpool Central University Research Ethics Committee C for healthy controls (Ref: 1672), and NRES Committee North West (Greater Manchester West, UK) for people with RA (Ref: 11/NW/0206). All participants gave written, informed consent in accordance with the declaration of Helsinki. All patients with rheumatoid arthritis fulfilled the American College of Rheumatology 2010 criteria for the diagnosis of RA. Disease activity was recorded using DAS28 score and improvements in disease activity measured using EULAR criteria [24]. Th early RA (ERA) cohort comprised newly-diagnosed patients prior to commencing disease-modifying anti-rheumatic drug (DMARD) therapy. Severe RA (SRA) patients were people with a DAS28 score ≥5.1 recruited prior to commencing anti-TNF (TNFi therapy). Healthy controls were recruited from staff at the University of Liverpool. All participants were over the age of 18 years and free of infection.

### 2.2 Neutrophil isolation and culture

Neutrophils were isolated from heparinised peripheral blood using Hetasep and Ficoll Paque as previously described [25]. Contaminating erythrocytes were lysed using ammonium chloride lysis buffer. Neutrophils were resuspended in RPMI 1640 media (Life Technologies) containing L-glutamine (2mM) and Hepes (25mM) at a concentration of 5x10^6^/mL unless otherwise stated. Neutrophil purity was routinely >97% and viability >98%.

### 2.3 RNA isolation

RNA was isolated from 10^7^ neutrophils using an optimised Trizol-chloroform protocol (Life Technologies) [26], precipitated in isopropanol and cleaned using the Qiagen RNeasy (mRNA) or miRNeasy (miRNA) kit including a DNase digestion step. RNA was snap-frozen in liquid nitrogen and stored at -80°C. Total RNA concentration and integrity were assessed using the Agilent 2100 Bioanalyser RNA Nano or Pico chip. RNA integrity was routinely ≥ 7.0.

### 2.4 mRNA sequencing

Total RNA was enriched for mRNA using poly-A selection. For cohort 1, 50 base pair single-end read libraries were sequenced on the Illumina HiSeq 2000 platform. For cohort 2, 100bp paired-end read libraries were sequenced on the DNBseq platform. Reads were mapped to the human genome (hg38) using HISAT2 [27]. Read counts were generated using featureCounts which is part of the Rsubread package (v2.0.1) [28] for R (v4.0.2) [29]. Statistical analysis of gene counts was carried out using edgeR (v3.28.1) [30] and limma [31] applying TMM normalisation and a false-discovery rate correction to p-values. Batch variation was removed using SVA [32].

### 2.5 miRNA sequencing

Total RNA was size selected (60-300nt), de-capped and sequenced on the Illumina NovaSeq platform. Micro RNAs reads were processed with miRge3.0 [33] which aligned reads to the human miRbase database (version 22) [34] with Bowtie [35]. Statistical analysis of miRNAs was performed using DESeq2 (contrasts RA vs HC) [36] in R (v4.0.2) [29], applying a Benjamini-Hochberg (BH) false-discovery rate correction to p-values.

### 2.6 Bioinformatics and statistical analysis

Gene ontology enrichment analysis of genes with 1.5-fold increased expression between RA vs HC was carried out using R package clusterProfiler against the genome wide annotation for human (org.Hs.eg.db) [37, 38] and included over representation searches in the Reactome database [39]. Gene set enrichment analysis was carried out in R using the CRAN package tmod [40] and based on a modular framework for classifying blood genomics studies [41]. Gene expression network analysis was carried out ARACNE2 [42] filtering on a mutual information (MI) threshold of 0.5 (p=10^-20^). Gene networks were visualised using Cytoscape [43] using Glay clustering within the clustermaker2 package [44]. Genes within each cluster were analysed for over representation of Gene Ontology categories using BINGO [45]. Functional enrichment analysis of genes with increased or decreased expression between RA vs HC, and miRNA:mRNA target enrichment analysis, was complemented with Ingenuity Pathway Analysis (https://digitalinsights.qiagen.com/IPA) [46] applying a 1.5-fold change in gene expression cut-off and a BH correction to p-values for canonical pathway analysis, using the Ingenuity Knowledge Base as background. Gene set variation analysis (GSVA) was performed using the GSVA package [47] against the Molecular signatures database (MSigDB) [48] in October 2023. Datasets used were C5 Gene Ontology Biological Processes (GOBP) and Human Phenotype Ontology (HPO), C7 immunologic signatures, C8 cell type signature gene sets.

Differential gene expression analysis of transcriptomics data is detailed above. Statistical analysis of experimental data was performed using R (v4.0.2) [29]. The Shapiro-Wilk test was complemented with QQ plot analysis to determine normality. Then univariate analysis was carried out by the Student’s t-test or Wilcoxon test as appropriate. Correlation test was performed in R (v4.0.2) and a significant correlation considered when the p-value post false discovery adjustment (Benjamini-Hochberg) was <0.05.

### 2.7 Protein extraction and Western blotting

Neutrophil proteins were extracted from Trizol lysates using manufacturer’s instructions into Laemmli buffer containing 1:1 3% SDS and 9M urea without bromophenol blue. Protein concentrations were measured using the BCA assay (Pierce) and normalised across samples before loading onto SDS-PAGE gel. Protein samples were separated by SDS-PAGE using a 12% gel and transferred onto PVDF membrane (Merck). Primary antibodies were: catalase (1:1,000 Abcam), glutathione peroxidase (1:1,000 Abcam) and GAPDH (1:1,000, Merck). Secondary antibodies were anti-rabbit IgG-specific HRP-linked (1:10,000, GE Healthcare) and anti-mouse IgG specific HRP-linked (1:10,000, Abcam). Bound antibodies were detected using the ECL system (Merck) and carefully exposed film to avoid saturation.

### 2.8 Enzyme-linked immunosorbent assays

Serum samples were collected using Z-clot serum vacutainers, and aliquots snap frozen in liquid nitrogen before storing at -80°C. Sera were clarified by centrifugation at 1000g for 15 min prior to performing ELISA. Interferon-α concentration was measured in serum using the VeriKine high-sensitivity interferon-alpha (all subtype) ELISA Kit (PBL assay science) using manufacturer’s instructions. The presence of NETs in sera was measured using the capture antibody from the human myeloperoxidase ELISA Kit (Abcam) and the anti-DNA detection antibody from the Cell Death Detection ELISAplus (Merck). The standard curve was comprised of NETs produced *in vitro* by human neutrophils in response to PMA, as previously described [49, 50]. ELISA analyte concentrations were determined by 4-PL regression with reference to the assay-specific standard curve.

### 2.9 Enzyme activity assays

Superoxide dismutase (SOD) activity was measured using the SOD activity colorimetric assay (Abcam). Catalase (CAT) activity was measured using the catalase activity fluorometric assay (Abcam). Glutathione peroxidase (GPx) activity was measured using the glutathione peroxidase activity colorimetric assay (Abcam). For all assays, neutrophils (2x10^6^) were washed with ice-cold PBS, centrifuged at 1,000g for 3 min, and pellets lysed in 200μL ice cold assay buffer. Cell lysates were centrifuged at 10,000g for 15 min at 4°C, and supernatant pipetted into a clean tube before snap freezing in liquid nitrogen and storing at -80°C prior to analysis. Enzyme activity assays were performed using manufacturer instructions and enzyme activity was calculated from either a standard curve (GPx and CAT assays) or expressed as a percentage inhibition rate based on positive and negative control wells (SOD assay) as described in the manufacturer instructions.

## 3. RESULTS

### 3.1 Neutrophil gene expression in severe RA is driven by interferons

In our previous study we identified that peripheral blood RA neutrophils have a different gene expression profile from healthy controls, driven typically by either a Type-I interferon-induced signature or expression of genes relating to neutrophil granule proteases [15]. For this study we recruited a second, larger, cohort of Biologics-naïve RA patients with high disease activity (DAS28 > 5.1), termed Severe RA (SRA). We combined our two cohorts of Severe RA patients together (n=65 in total) and compared gene expression to healthy control (HC, n=11) neutrophil transcriptomes. We determined that 289 genes were significantly different between SRA and HC neutrophils (FDR adj p<0.05, Figure 1A). The top 1% of significantly different genes were all down-regulated in SRA neutrophils compared to HC (i.e. higher in HC neutrophils, Figure 1B). Functional over-representation analysis of these genes using the Reactome database against total resources for *Homo sapiens* identified enrichment of signalling pathways for ‘Regulation of TP53 activity through association with co-factors’ and ‘Regulation of NFE2L2 gene expression’ (FDR adj p< 0.05). SRA neutrophils showed high heterogeneity in the top 1% of up-regulated genes (Figure 1B), with distinct clusters of SRA patients with “high” or “low” interferon-response gene expression. Reactome analysis identified significant enrichment of signalling pathways from ‘Interferon alpha/beta’, ‘interferon gamma’ and ‘OAS antiviral response’ (all FDR adj p<0.05).

**Figure 1.**
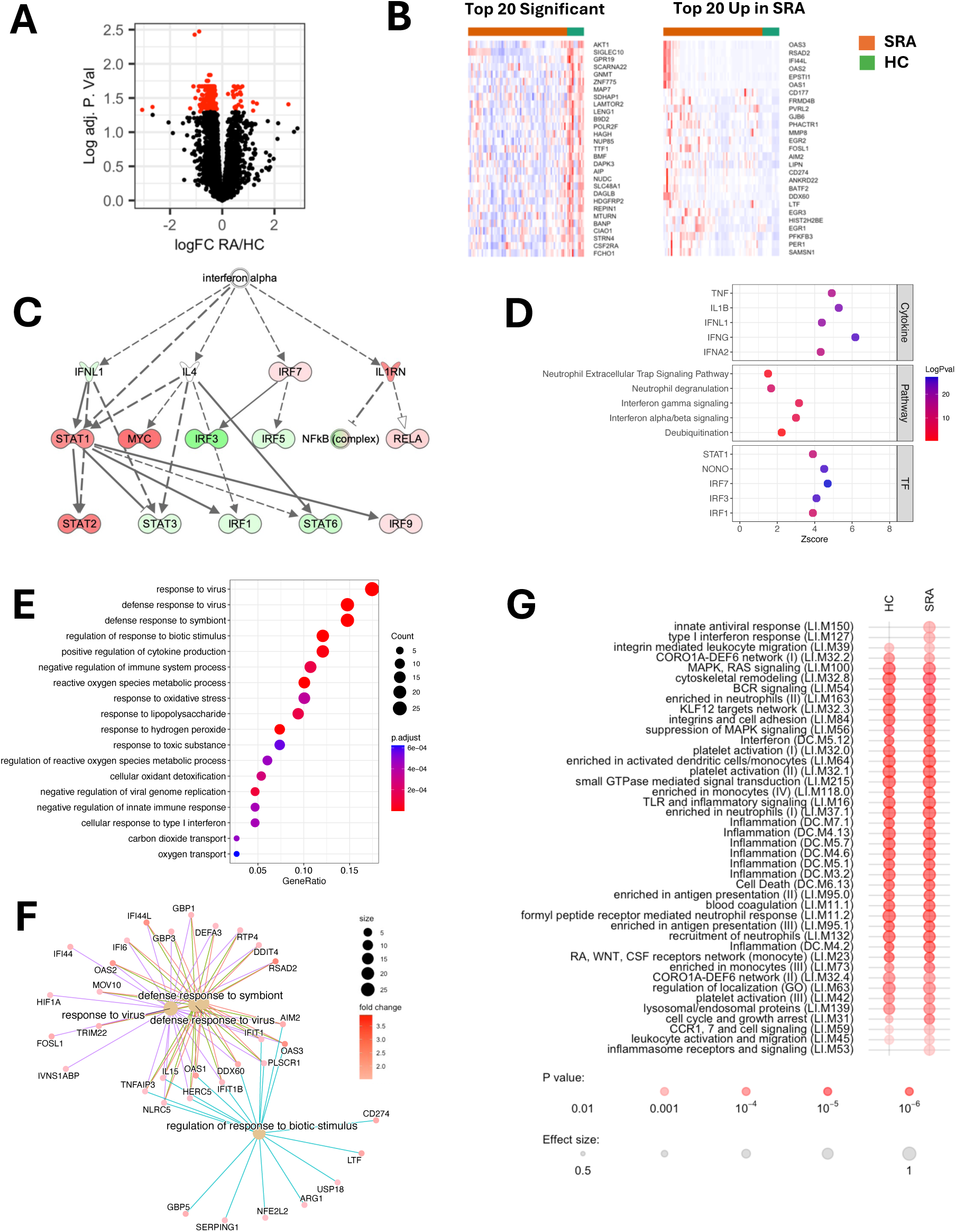
Transcriptomic analysis of severe RA (SRA) neutrophils. (A) Volcano plot showing genes highlighted in red up and down-regulated in SRA compared to HC (adj p<0.05). (B) Heatmaps showing top 1% significant and top 1% up-regulated genes in SRA compared to HC. Orange bar indicates SRA samples and green bar indicates HC samples. (C) IPA prediction of interferon signalling genes upregulated (red) in SRA. Green indicates down-regulated gene. (D) Summary of IPA upstream analysis of cytokines, canonical pathways and transcription factors (TF) activated in SRA neutrophils. (E) Gene ontology analysis of genes up-regulated 1.5-fold in SRA neutrophils. (F) Plot of genes common to top signalling pathways relating to defence response to viral infections. (G) Modular enrichment analysis (tmod) of genes expressed in SRA and HC neutrophils (AUC >0.8).

In order to investigate the altered transcriptome of SRA neutrophils further we used IPA to predict major signalling pathways regulating SRA neutrophil gene expression. As previously described, an interferon alpha regulated network of genes was the highest predicted interaction network by IPA (Figure 1C, adj p<0.01). Interferon gamma and interferon alpha signalling were the most highly predicted canonical pathways activated in SRA neutrophils (Figure 1D, adj p<0.01). Other pathways significantly activated were neutrophil degranulation, neutrophil extracellular trap signalling and deubiquitination (Figure 1D, adj. p<0.01). Upstream regulator cytokine analysis predicted regulation of SRA neutrophil gene expression by interferons (IFNA1, IFNG, IFNL1), IL-1β and TNFα (Figure 1D, adj p<0.01, Supplementary Table 1). Predicted transcription factor activation included IRF1, IRF3 and IRF7 as well as STAT1 and NONO (Figure 1D, adj p<0.01, Supplementary Table 1). We complemented IPA analysis with GO analysis accessed through the R package clusterProfiler. We confirmed HC vs SRA differences, relating to interferon signalling, innate immune activation and response to oxidative stress and reactive oxygen species (Figure 1E). Many of the up-regulated genes in SRA were common to several gene ontologies including the ‘defence response to a virus’ and ‘response to a biotic stimulus’, and the relationships between these 33 genes were visualised in a cnetplot (Figure 1F). Many of these genes have been described as type-I interferon response genes (e.g. IFI44L, IFI6, OAS1, OAS2, OAS3 and RSAD2), expression of which was confirmed in SRA neutrophils by qPCR in our previous studies [14, 15].

We next used blood transcriptional modular enrichment analysis (R package tmod) to identify gene expression modules enriched in SRA and HC neutrophils across our two separate cohorts. This modular analysis is based on signalling modules compiled from analysis of blood transcriptomes across a number of inflammatory conditions [41]. Once again modules relating to interferon response and inflammasome activation were enriched in SRA neutrophils compared to HC. In addition, this analysis identified enhanced expression of genes regulating the cell cycle response, which we previously showed was enhanced in RA low-density neutrophils [51] (Figure 1G, Supplementary Figure 1A). Preliminary proteomics analysis of SRA neutrophils (n=3) confirmed the elevation of the type-I interferon response at the protein level, with expression of several type-I interferon response proteins being >2-fold higher in SRA neutrophils compared to healthy controls (data not shown).

### 3.2 Neutrophil gene expression in treatment-naïve RA is similar to severe RA

We next sought to determine whether RA neutrophil gene expression changes during the course of the disease. We therefore isolated peripheral blood neutrophils from a cohort of newly diagnosed, DMARD-naïve patients (Early RA cohort, ERA, n=13). Only one gene (MAPK14) was significantly different between ERA patients and HC (Figure 2A, adj p<0.05). IPA revealed once again that the pathways up-regulated in Early RA (ERA) neutrophils were interferon alpha/beta signalling, interferon gamma signalling and neutrophil degranulation as well as pyroptosis signalling and ISGylation signalling (Figure 2B, adj. p<0.01). Upstream cytokine regulators of ERA gene expression were predicted to be interferons alpha, beta, gamma and lambda and IL-27 (Figure 2B, adj p<0.01, Supplementary Table 2). Similarly to SRA, the transcription factors predicted to be activated were IRF1, IRF3 IRF7, NONO and PML (Figure 2B, adj p<0.01, Supplementary Table 2). Gene ontology analysis of ERA neutrophil gene expression was surprisingly similar to SRA in that a network of 38 up- regulated genes were predicted to be enhancing the defence response to a virus and type-I interferon signalling (Figure 2C,E, adj p<0.01). Expression of interferon response genes IFI44L, RSAD2 and FCGR1A was similar in ERA and SRA, both of which were higher than in HC (Figure 2D). Modular gene set enrichment analysis with tmod showed comparable module enrichment across both ERA and SRA patient cohorts (Figure 2F), with more significant enrichment of gene modules relating to the innate antiviral response, the antiviral interferon signature and RIG-1 like receptor signalling in SRA (Figure 2F, Supplementary Figure 2). This led us to conclude that the neutrophil gene expression signature is enhanced but not significantly changed when RA disease activity does not resolve with DMARDs.

**Figure 2.**
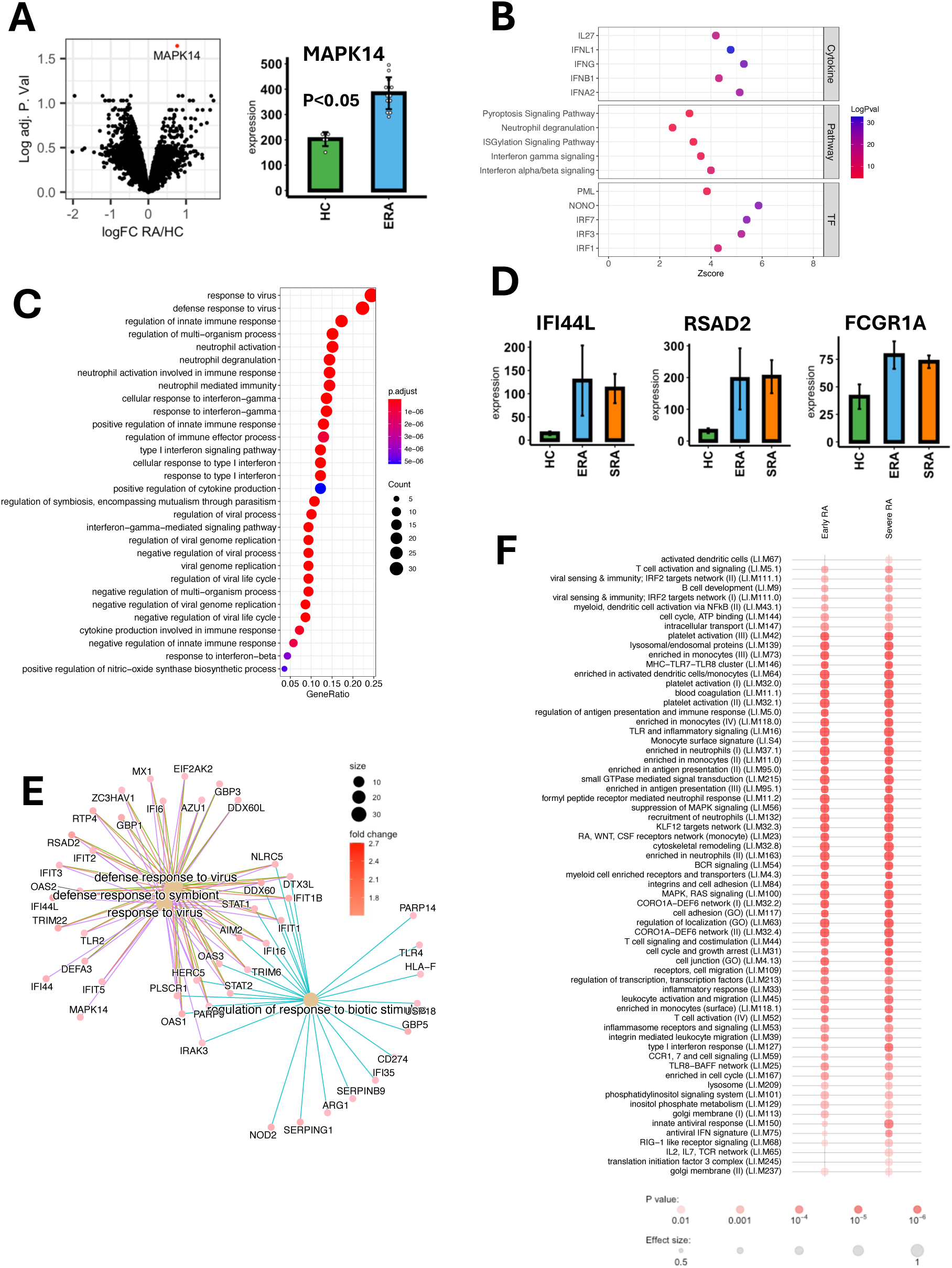
Transcriptomic analysis of early RA (ERA) neutrophils. (A) Volcano plot showing single gene highlighted in red as being significantly up-regulated in ERA compared to HC (adj p<0.05). Box- plot showing expression of MAPK14. (B) Summary of IPA upstream analysis of cytokines, canonical pathways and transcription factors (TF) activated in ERA neutrophils. (C) Gene ontology analysis of genes up-regulated 1.5-fold in ERA neutrophils. (D) Box plots showing expression of interferon- response genes IFI44L, RSAD2 and FCGR1A in HC, ERA and SRA neutrophils. (E) Plot of genes common to top signalling pathways relating to defence response to viral infections. (F) Modular enrichment analysis (tmod) of genes expressed in ERA and SRA neutrophils (AUC >0.8).

### 3.3 Gene networks regulate RA neutrophil gene expression

We next wanted to further investigate the regulation of gene networks in SRA neutrophils. To do this we used ARACNE2 to reconstruct the gene expression networks in SRA neutrophils, applying a mutual information (MI) threshold of 0.5 (p<10^-20^). Modules were reconstructed using Cytoscape and clustermaker [43, 44] and analysed for gene ontology overrepresentation using the BINGO app [45] and for canonical pathway enrichment using IPA. We identified six major modules of gene expression networks in SRA neutrophils (Figure 3, Supplementary Table 3). Module 1 (M1) related to metabolism and transcription, incorporating essential cell functions such as regulation of transcription, protein modification and localisation, metabolic processing of nucleosides and amino acids, and cellular response to stress. M2 was regulated by integrins and cytokine receptors including IL-6 and IL-6 signalling, NF-κB, NFAT and HMGB1 signalling and signalling by Rho GTPases. M3 regulated kinase activation including AMPK, SAPK/JNK, Cdc42 and NF-κB as well as 3-phosphoinositide biosynthesis. M4 was a much smaller network of genes regulated by interferon and toll-like receptor (TLR) receptor signalling. M5 was another module regulating gene expression this time incorporating phospholipase C (PLC) and apoptosis signalling as well as gene translation and metabolism of proteins. Finally M6 contained a gene network regulating carbohydrate and amino acid metabolism.

**Figure 3.**
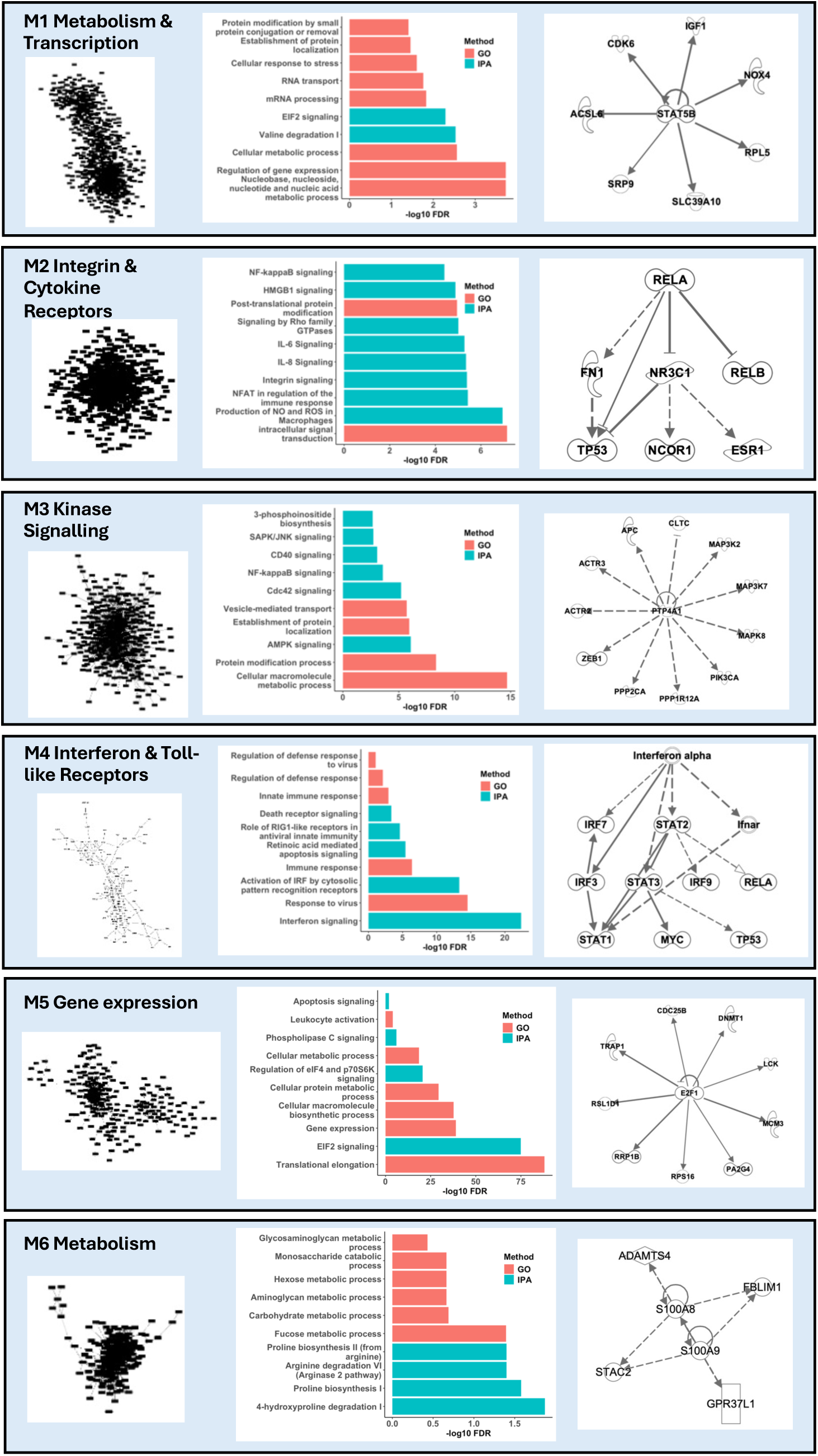
Modular gene network analysis identified six major gene networks in SRA neutrophils. Each panel shows reconstructed network numbered Mx, with bar chart showing gene ontologies (pink) and IPA canonical pathways (blue) activated within the gene network. Right panel in each module indicates up-steam regulator ‘hub’ genes within each module.

### 3.4 NET debris is elevated in RA sera

Our next aim was to validate the bioinformatics predictions and determine the factor(s) regulating neutrophil gene expression in SRA. We attempted to measure the level of interferon-alpha (IFN-α) in SRA sera collected at the same time as the neutrophils we subjected to RNAseq. We were unable to detect any IFN-α in SRA or HC sera using commercial ELISA kits (data not shown). However, using a high-sensitivity IFN-α (all subtype) ELISA we detected very low concentrations of IFN-α in 12 out of 24 SRA sera assayed (mean 0.8pg/mL) compared to 2 out of 10 HC sera (mean 0.47pg/mL). This difference was not statistically significant (Figure 4A, p=0.197) and the concentrations of IFN-α in SRA sera did not correlate with expression of interferon-response genes (Figure 4B, p=0.25). We also performed a custom sandwich ELISA on SRA and HC sera to measure the presence of NET debris (MPO:DNA complexes), and this identified significantly higher levels of NET debris in RA sera compared to healthy controls (Figure 4C, p<0.05).

**Figure 4.**
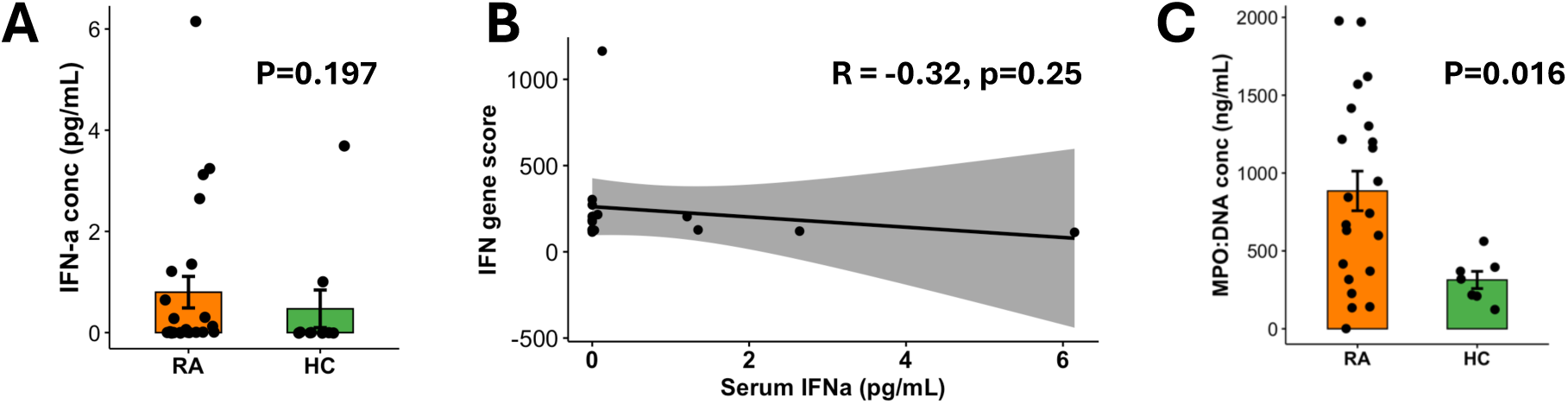
Serum inflammatory markers in people with SRA. (A) Serum interferon-alpha (all types) concentrations measured by high-sensitivity ELISA in SRA (orange) and HC (green) sera. (B) Correlation of interferon-alpha serum concentrations with average expression of interferon-response genes (IFN gene score) in neutrophils. (C) Concentrations of NET debris (DNA:MPO complexes) in SRA (orange) and HC (green) sera (p<0.05).

### 3.5 Antioxidant proteins are lower in RA neutrophils

We validated the role of oxidative stress in RA neutrophils, as this was identified by our gene ontology (Figure 1E). Western blot analysis of neutrophil protein lysates showed lower expression of catalase (Figure 5A,B, p<0.05) and glutathione peroxidase (Figure 5A,B, p<0.06) in SRA neutrophils compared to HC. Enzyme activity assay of neutrophil cell lysates identified that GPx activity was lower in SRA neutrophils (Figure 5C, p<0.01) and that SOD activity was elevated in SRA but not to a statistically significant level. We were not able to detect biologically active catalase using either a colorimetric or fluorometric enzyme activity assay (data not shown).

**Figure 5.**
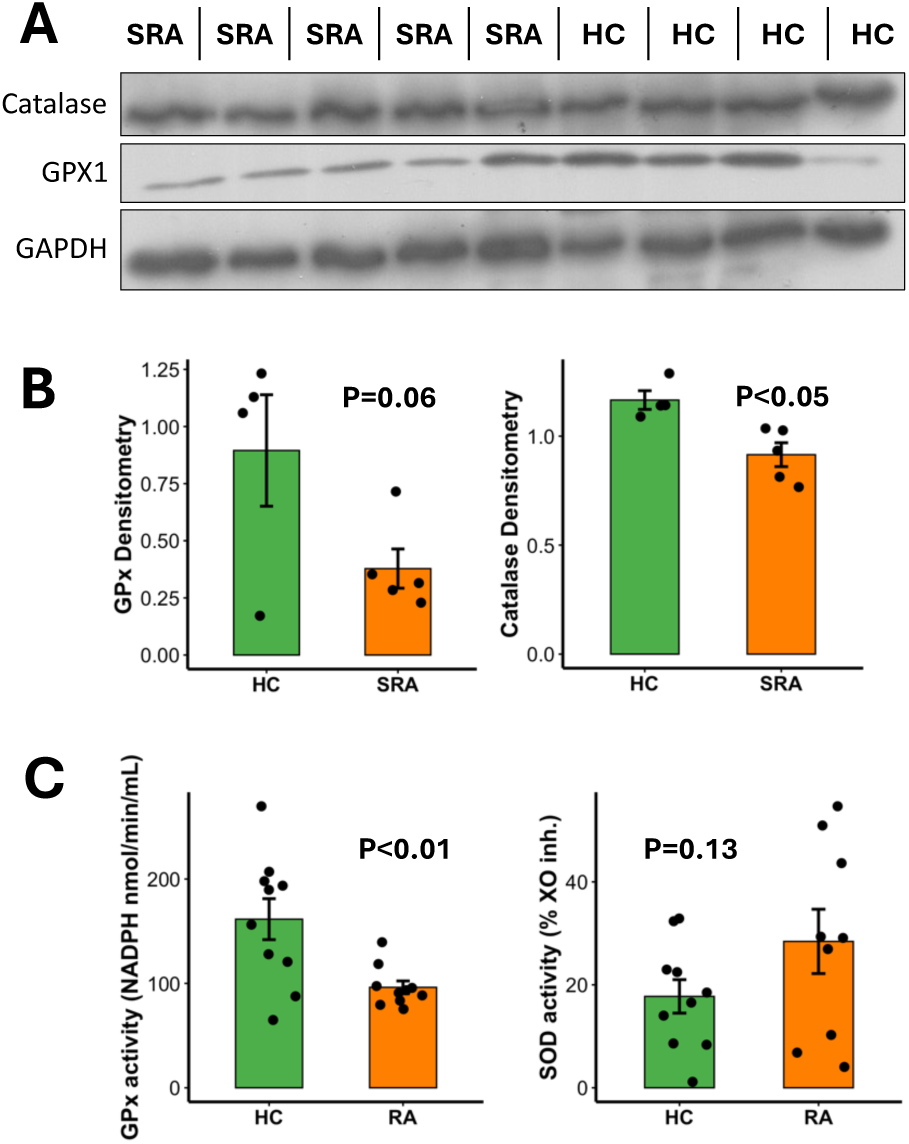
Anti-oxidant enzyme expression and activity in RA neutrophils. (A) Protein levels measured by western blotting of CAT and GPx. (B) Western blot of protein lysates from SRA (n=5) and HC (n=4). (C) Enzyme activity levels of CAT, GPx and SOD in SRA (orange) and HC (green) neutrophils.

### 3.6 MicroRNA regulation of RA neutrophil gene expression

We next wanted to determine the effect of micro RNAs on neutrophil mRNA expression in SRA. IPA upstream regulator analysis predicted that the type-I interferon response gene signature was regulated by microRNAs miR-183 (Figure 6A, p=4.64x10^-9^, Supplementary Table 4) and miR-96 (Figure 6A, p=8.23x10^-9^, Supplementary Table 4) [52]. We therefore performed microRNA sequencing on duplicate samples from a subset of SRA (n=6) and HC (n=5) neutrophils. We observed increased expression of miR-183 (Figure 6B, p<0.01, adj p=0.1) and miR-96 (Figure 6B, p<0.05, adj p=0.24) in SRA neutrophils however neither reached significance after p-value adjustment. There was clear separation of SRA and HC miRNAseq samples by principal component analysis (PCA, Figure 6C) and we identified 20 miRNAs with significantly different expression in SRA neutrophils (Figure 6D, adj p<0.05). Using the mRNA:miRNA filter function in IPA, we confirmed that interferon alpha/beta signalling was the most highly predicted canonical signalling pathway activated in SRA neutrophils (adj. p = 1.17x10^-6^), with microRNAs miR-182-5p, miR-183 and miR-96 predicted to be regulating interferon-response gene expression (Figure 6E, all p<0.01, Supplementary Table 5). The interferon alpha receptor was predicted to be activated (p=7.28x10^-14^) as were the interferon activated transcription factors IRF1, IRF3, ORF7 and STAT1 (p<0.01, Supplementary Table 5). In addition, IPA predicted the activation of key regulators of mRNA and miRNA expression AGO2 (p=1.97x10^-45^) and DICER1 (p=3.33x10^-15^).

**Figure 6.**
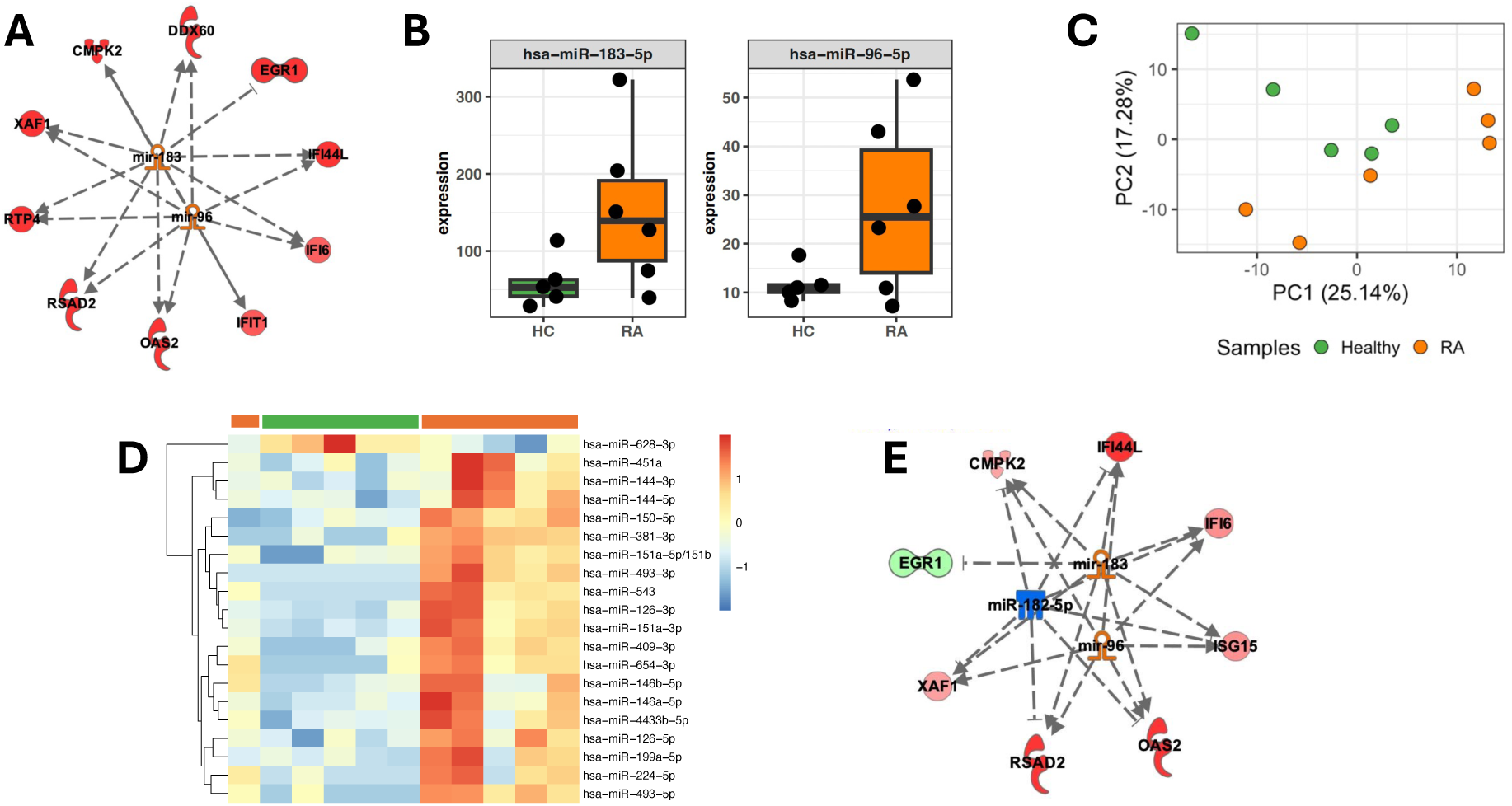
microRNA expression in SRA neutrophils. (A) Predicted miRNA expression in SRA neutrophils from IPA analysis of transcriptomics data showing hub miRNAs miR-183 and miR-96 and expression of interferon- response genes (up-regulated in red). (B) Measured expression of miR-183-5p and miR-96-5p in paired SRA (orange) and HC (green) neutrophils (p<0.05, adj p >0.05). (C) PCA analysis of HC and SRA miRNAseq data. (D) Heatmap of significantly expressed miRNAs from SRA (orange) and HC (green) neutrophils measured by miRNAseq (adj p<0.05). Colour range is from logFC of -2 to +2, chosen because it is representative of the distribution of the expression data without being dominated by outliers. (E) Network reconstruction using IPA of interferon-response genes and miRNAs expressed in RA neutrophils (red = up-regulated mRNA, green = down-regulated mRNA, orange = up-regulated miRNA, blue = down-regulated miRNA).

### 3.7 Identification of neutrophil subset gene expression signatures

Recent insight from single cell RNA studies of mouse and human neutrophils has identified subsets of blood neutrophils which can be defined by distinct expression of genes that are not necessarily expressed at the protein level. A study by Wigerblad et al. [53] identified the trajectory of healthy human neutrophils from immature (Nh0) to intermediate (Nh1) stages, which then diverge to either Nh2 or Nh3 mature polarised neutrophils, based on the expression of distinct transcription factor genes. The Nh3 gene cluster, representing around 7% of blood neutrophils in healthy donors, includes a number of interferon-response genes including IFI6, IFIT1 and RSAD2. Using the published list of Nh marker genes [53], we compared the level of expression of genes representing each neutrophil subtype in our bulk RNAseq data from SRA and HC neutrophils. We found that two Nh0 genes, LCP1 and TLR4, were significantly higher in SRA neutrophils compared to HC (Figure 7A, adj. p<0.05). Three Nh1 marker genes, SIGLEC10, SQSTM1 and SEC14L1, were significantly lower in SRA neutrophils (Figure 7A, adj. p<0.05). Nineteen genes marking the Nh2 cluster were significantly different in SRA neutrophils. Of these, fifteen were lower in SRA neutrophils including CSF2RA, ATXN2 and BANP (Figure 7A, adj. p<0.05). While there was no significant difference in the expression of Nh3 genes between the SRA and HC groups, the most upregulated genes based on fold change were the IFN-regulated genes RSAD2, OAS3 and IFI44L, due to the previously described IFN-high and IFN-low sub- populations of SRA patients (Figure 7B). We used IPA to predict which micro RNAs were regulating the expression of SRA genes from each Nh cluster. Elevated expression of Nh0 genes was predicted to be regulated by the decrease in expression of miR-146a-5p (adj p=8.03x10^-3^), Nh1 and Nh2 genes were down-regulated by increased expression of miR-155-5p (Nh1 adj p=1.13x10^-6^; Nh2 adj p=6.17x10^-5^), and Nh3 genes were predicted to be regulated by miR-183 (adj. p=1.45x10^-22^) and miR- 96 (adj. p=5.03x10^-22^, Figure 7C).

**Figure 7.**
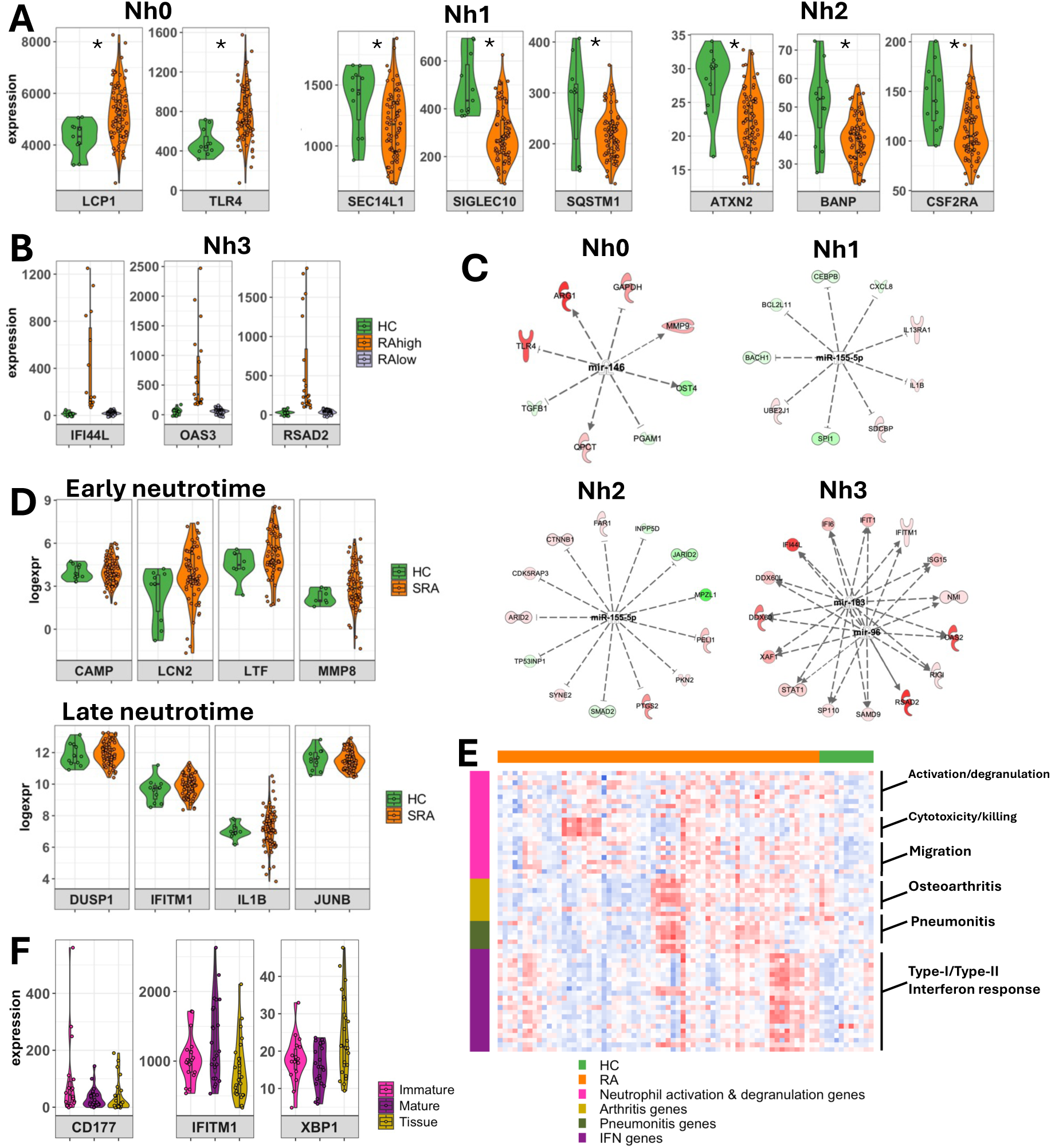
Identification of neutrophil subsets in RA neutrophils using publicly–available single-cell RNAseq datasets. (A) Expression levels of gene markers of human neutrophil (Nh) subsets in SRA (orange) and HC (green) neutrophils (Nh0, Nh1, Nh2, Nh3, * adj.p<0.05). (B) Nh3 subset characterized by RA interferon (IFN) high and low patients. (C) Predicted miRNA regulated gene networks for each Nh subset. (D) Expression of early and late “neutrotime” genes in SRA and HC neutrophils. (E) GSVA analysis of RA (orange) and HC (green) neutrophils identifies gene signatures relating to early neutrophil activation (pink), interferon genes (purple), arthritis genes (yellow) and systemic inflammation associated with tissue inflammation (pneumonitis genes (dark green)). (F) Expression levels of marker genes associated with polarization of early/immature neutrophils (CD177, pink), mature neutrophils (IFITM1, purple) and tissue-specific inflammation (XBP1, yellow) across the SRA cohort.

In another study, Grieshaber-Bouyer et al [54] performed single cell RNAseq analysis of mouse neutrophils, which identified sub-clusters of neutrophils with distinct gene expression profiles that track maturity along a trajectory from bone marrow into peripheral blood (termed “neutrotime”), with polarised states along neutrotime arising as a result of inflammatory stimulus and tissue-specific gene expression. Distinct neutrotime clusters were associated with gene expression patterns unique to both maturity state and the site of inflammation, which included KxB/N inflammatory arthritis model, IL-1β-induced pneumonitis and IL-1β-induced peritonitis. We therefore wanted to determine whether we could detect different clusters of neutrotime gene signatures amongst our bulk RNAseq datasets. First, we compared the expression of early neutrotime genes (CAMP, LCN2, LTF, MMP8) and late neutrotime genes (DUSP1, IFITM1, IL1B, JUNB) in our SRA and HC transcriptomes (Figure 7D). We found that whilst none of these genes had significantly different expression between the SRA and HC groups, a clear subset of SRA patients with high expression of early neutrotime genes was evident (Figure 7D), suggesting that the population of immature neutrophils may be increased in these individuals.

We next performed gene set variation analysis (GSVA) of our SRA and HC neutrophil transcriptomes against the molecular signatures database (MSigDB) including gene ontology sets and human phenotype signatures to investigate potential neutrotime signatures. This analysis allows to us to collapse our gene level information to relevant pathways or processes, in this case immune-based, while retaining sample-specific information. We found that the SRA transcriptomes had distinct phenotypes, relating to one of four molecular signatures: (1) neutrophil activation and degranulation, (2) interferon-regulated genes, (3) arthritis and (4) pneumonitis (Figure 7E). These broadly relate to gene markers associated with neutrotime polarisation states representing immature neutrophils (CD177), mature neutrophils (IFITM1), and tissue specific neutrophils (XBP1) (Fig 7F).

## 4. DISCUSSION

In this study we have generated transcriptomic data from early and severe RA and HC neutrophils to investigate the signalling pathways that contribute to neutrophil-driven inflammation in RA. Our analyses revealed a significant increase expression of genes of the Type-I (IFN-α/β) signalling pathway in neutrophils from a sub-population of people with early and severe DMARD-refractory RA. We identified a cluster of microRNAs predicted to regulate type-I interferon response gene expression. We also identified expression of marker genes associated with sub-populations of blood neutrophils previously identified in single-cell RNAseq studies that are predicted to be regulated by specific miRNAs [53, 54].

The most profound signature that characterised RA neutrophils was a type-I interferon induced signature. Interferons are a group of functionally related cytokines that are produced in response to viral infection [55]. Human neutrophils are highly responsive to interferons, which can alter the rate of apoptosis, enhance TNF-α-primed ROS production and augment cytokine and chemokine synthesis [25, 56]. Interferons signal through dedicated IFN-α/β or IFN-γ receptors which activate JAK-STAT signalling and IRF family transcription factors, and the small molecule JAK inhibitors baricitinib and tofacitinib can modulate RA neutrophil activation [56, 57]. Our analysis showed that interferon-induced gene expression was the most significant signature in SRA neutrophils, and this activation signature was also evident in DMARD-naïve ERA neutrophils. Interestingly, our analysis suggests that gene expression in ERA and SRA neutrophils is remarkably similar, possibly reflecting uncontrolled, systemic inflammation. All RA patients in our study had active inflammation, with DAS28 scores >3.2 in the ERA cohort, and >5.1 in the SRA cohort.

The interferon-response genes identified in SRA neutrophils play important roles in cellular inflammation and neutrophil activation. Oligoadenylate synthase (OAS) genes encode enzymes that sense exogenous nucleic acids and initiate antiviral pathways. High levels of OAS family genes are strongly associated with autoimmune diseases, including RA [58–60]. RSAD2 encodes an interferon-inducible antiviral protein of the S-adenosyl-L methionine enzyme superfamily that can regulate both immune cell proliferation and Th2 cell polarization via NF-κB, GATA3 and IL-4 [61]. CMPK2 is a mitochondrial enzyme crucial for maintaining mitochondrial DNA integrity. It is located on the short arm of chromosome two (2p), positioned inverted and adjacent to the RSAD2 gene [62]. Both these genes are co-transcribed upon the induction of interferon-induced signalling [63, 64]. Altered levels of CMPK2 have been reported to disrupt mitochondrial physiology and significantly de-regulate macrophage homeostasis. Constitutive overexpression of the CMPK2 in macrophages induces mitochondrial stress with marked depolarization of membrane potential, increases expression of pro-inflammatory genes (IL-8, IL-1β and TNF-α) and enhances ROS production. Interestingly, sustained CMPK2 expression results in a heightened glycolytic activity in macrophages [65] something that is also observed in RA neutrophils [57]. IFI44L typically modulates cellular environments to make them hostile to viruses, but dysregulation of this process may promote inflammation and targeting of self-tissues. High IFI44L levels correlate with RA disease activity [66], and may be a useful diagnostic tool to measure disease severity [67]. IFI16 is a sensor for exogenous DNA in macrophages, responsible for stimulating early innate immune responses and inflammation via the activation of inflammasomes. These cytoplasmic complexes are capable of processing and releasing pro-inflammatory IL-1β [68]. Furthermore, IFI16 activates stimulator of interferon genes (STING), consequently inducing NF-κB signalling in a cyclic GMP-AMP synthase (cGAS)-independent manner. As well as recognising microbial DNA, cGAS can bind self-DNA leaked from mitochondria or the nucleus effectively triggering extensive inflammatory responses [69]. Increased expression of cGAS-STING could therefore indicate a role for NETs in driving neutrophil activation and type-I interferon response genes in RA [70]. As well as detecting elevated NETs in SRA sera in this study, we recently identified a sub-population of DESpR positive, NETting neutrophils in whole blood smears from people with RA [23] confirming that NET production takes place in circulating RA blood as well as within synovial joints and tissue in RA [6, 9]. A recent clinical study also detected elevated NET debris (DNA:elastase, and DNA:histone H3 complexes) in RA plasma, although this did not reach statistical significance or correlate with disease activity [71].

Our bioinformatics analyses identified a key role of miRNAs miR-182-5p, miR-183 and miR-96 in regulating RA neutrophil gene expression. These miRNAs exist in a cluster known as miR-183C [72]. This cluster is highly conserved and is transcribed as a polycistronic miRNA cluster, known to promote cancer development. These miRNAs possess almost identical seed sequences, and this similarity enables them to act co-operatively [73]. The miR-183C cluster positively regulates type 1 IFN production by inhibiting negative regulators of the antiviral response. Specifically, miR-183C targets and silences PP2A, a known inhibitor of transcription factors STAT1 and IRF3 [74, 75] leading to enhanced IRF3 signalling in the initiation of type 1 IFN production [68]. The miR-183C cluster is also known to play roles in immunity and autoimmunity, with its dysregulation reported in various autoimmune disorders including multiple sclerosis, systemic lupus erythematosus (SLE), and ocular autoimmune disorders [76]. In murine models of SLE and multiple sclerosis inhibition of this cluster *in vivo* has yielded positive therapeutic outcomes, with a significant attenuation of disease symptoms [76–78]. This has led to the prospect of developing miR-183C miRNA-based therapies for targeting autoimmune diseases.

By comparing our bulk RNAseq data to single cell RNAseq datasets we were able to identify gene signatures relating to neutrophil subsets. Two recent publications have shed new insight onto the complexity and heterogeneity of the blood neutrophil population. A single-cell RNAseq study of mouse neutrophils identified a transcriptional trajectory program during neutrophil development, maturation and migration into tissues that was termed “neutrotime” [54]. Each maturation step could be identified by marker genes, and indeed we detected these gene signatures in our SRA neutrophils. We found that we could identify a clear subset of patients who had neutrophils expressing either early neutrotime or late neutrotime marker genes. The early neutrotime genes code for neutrophil granule proteins such as lactoferrin (LTF) and lipocalin-2 (LCN), which we have previously reported to be highly expressed in a subset of RA patients, and in RA low-density granulocytes [14, 51]. These genes are normally silenced during neutrophil development before mature neutrophils exit the bone marrow and enter circulation [79], and this could indicate high levels of inflammation and increased granulopoiesis in these individuals. We were also able to compare our bulk RNAseq data to a human neutrophil single-cell RNAseq study [53] which again identified distinct maturation subsets of blood neutrophils from healthy individuals. Our analysis showed that SRA neutrophils expressed genes for all Nh subsets identified in this study, and in addition we found a sub-set of SRA patients with increased expression of Nh3 interferon marker genes. We did not identify elevated serum levels of interferon-alpha in the patients with high expression of interferon-response genes raising the question as to whether it is indeed interferons that are activating interferon-response genes in SRA, or whether it is some other agonist such as viral or NET-derived DNA via the cGAS-STING system. We also predicted regulation of neutrophil Nh subsets by microRNAs miR-146a-5p and miR-155-5p. Decreased expression of miR-146a-5p has been shown to increase expression of genes such as TRAF6 and IRAK1, which are involved in activating NF-κB signalling and promoting inflammation [80], whereas in mice, miR-155 has been identified as a key regulator of PAD4 activation and NET release [81].

As well as altered gene expression, we identified an altered, activated phenotype in our SRA neutrophil cohort. Our gene over representation analysis identified several ontologies relating to the response to oxidative stress, hydrogen peroxide and reactive oxygen species in SRA neutrophils, and Reactome analysis identified significantly lower expression of genes relating to the regulation of NFE2L2 gene expression. The NFE2L2 gene encodes the transcription factor Nrf2, a key regulator of expression of antioxidant genes. Enzyme activity assays revealed higher levels of SOD activity in some, but not all, RA patients compared to healthy controls. This enzyme is involved in the reduction of the superoxide radical to hydrogen peroxide for example during the neutrophil respiratory burst [11]. In contrast, we detected significantly lower levels of GPx activity in RA neutrophils. This enzyme plays an important role in detoxification of hydrogen peroxide by reducing it to water. Taken together, this evidence would suggest that RA neutrophils have a dysregulation of hydrogen peroxide detoxification which would cause oxidative stress leading to the production of hypochlorous acid through the action of myeloperoxidase. As well as inducing damage to tissues, DNA and protein, low levels of hypochlorous acid can rapidly de-activate glutathione peroxidase [82] suggesting a possible intracellular mechanism for the results observed.

We recognise several limitations to our study. Our blood samples were non-fasted and collected from multiple clinics at different times of day. While there is mounting evidence of the effect of metabolism and circadian rhythms on neutrophil activation and response to infection [83] [57], we did not identify any gene expression patterns relating to the time of blood collection in our data (data not shown). In addition, we measured RNA expression in blood neutrophils isolated by density-centrifugation, meaning we likely lost low-density neutrophils and, potentially, fragile NETting neutrophils during the isolation process [23]. We were also unable to obtain neutrophils from the synovial joint as part of this study, although we have previously reported the distinct gene expression profile of RA synovial fluid neutrophils [9]. Despite these limitations, we believe our study sheds important new insight onto the complexity and heterogeneity of RA blood neutrophil gene expression and provides a potential mechanism for regulation of neutrophil subsets, and the interferon-response gene signature detected in a sub-set of people with RA, by microRNAs.

## 5. CONCLUSION

In conclusion, we have described a complex and heterogeneous neutrophil transcriptome across people with RA. These gene expression profiles are conserved across patients with early-RA and severe, DMARD-refractory RA. The major gene signature within the cohort as a whole is an interferon-response gene signature, and we propose this is regulated by the microRNA hub of miR-182, miR-183, miR-96 (miR-183C cluster). Interferon alpha was not significantly elevated in RA sera, but we have detected increased levels of circulating NET debris which may activate interferon-gene expression via the DNA-sensing genes (cGAS-STING). The heterogeneous transcriptomes measured over the population of people with RA can be broadly compared to single RNAseq datasets which identify hub genes regulating polarised states of neutrophil subsets. These neutrophil subsets are predicted to be regulated by different microRNAs including miR-146a5p, miR-155-5p, and the miR-183C cluster. We believe our study provides valuable insight into the activation of neutrophils in the blood of people with RA and defines the heterogeneity of neutrophil subsets in people with RA warranting further investigation by single-cell sequencing of RA blood neutrophil populations.

## 6. ABBREVIATIONS

ACPA,: anti-citrullinated protein antibodies
CAT,: catalase
DAS28,: 28-joint disease activity score
DEspR,: dual endothelin-1/VEGF signal peptide receptor
DMARD,: disease-modifying anti-rheumatic drug
ERA,: early rheumatoid arthritis
GPx,: glutathione peroxidase
GSVA,: gene set variation analysis
HC,: healthy control
IFN,: interferon miRNA, microRNA
NET,: neutrophil extracellular trap
PLS-DA,: partial least squares discriminant analysis
RA,: rheumatoid arthritis
ROS,: reactive oxygen species
SOD,: superoxide dismutase
SRA,: severe rheumatoid arthritis

## Data Availability

All data produced in the present study are available upon reasonable request to the authors

## ACKNOWLEDGEMENTS

We would like to thank clinical colleagues at hospitals in the Merseyside region for support with participant recruitment and sample collection. We would also like to thank Miss Robyn Campbell for technical support.

## 8. AUTHOR CONTRIBUTIONS

MFA formal analysis, methodology, visualisation, writing – review and editing

GAA formal analysis, writing – review and editing

JAB formal analysis, writing – original draft

IK formal analysis, writing – original draft

AS formal analysis, validation

AC formal analysis, validation

SH formal analysis, methodology, writing – review and editing

PA, methodology, visualisation

ECG, methodology, visualisation, writing – review and editing

HLW conceptualisation, data curation, formal analysis, funding acquisition, investigation, methodology, project administration, supervision, validation, visualisation, writing – original draft, writing – review and editing

## REFERENCES

[1] M. Fresneda Alarcon, Z. McLaren, H. L. Wright. Neutrophils in the Pathogenesis of Rheumatoid Arthritis and Systemic Lupus Erythematosus: Same Foe Different M.O. Front Immunol, 2021;12:649693.

[2] H. L. Wright, R. J. Moots, R. C. Bucknall, S. W. Edwards. Neutrophil function in inflammation and inflammatory diseases. Rheumatology (Oxford), 2010;49:1618–31.

[3] H. L. Wright, R. J. Moots, S. W. Edwards. The multifactorial role of neutrophils in rheumatoid arthritis. Nature reviews Rheumatology, 2014;10:593–601.

[4] N. Thieblemont, H. L. Wright, S. W. Edwards, V. Witko-Sarsat. Human neutrophils in auto- immunity. Seminars in immunology, 2016;28:159–73.

[5] J. A. Quayle, F. Watson, R. C. Bucknall, S. W. Edwards. Neutrophils from the synovial fluid of patients with rheumatoid arthritis express the high affinity immunoglobulin G receptor, Fc gamma RI (CD64): role of immune complexes and cytokines in induction of receptor expression. Immunology, 1997;91:266–73.

[6] J. Spengler, B. Lugonja, A. J. Ytterberg, R. A. Zubarev, A. J. Creese, M. J. Pearson et al. Release of Active Peptidyl Arginine Deiminases by Neutrophils Can Explain Production of Extracellular Citrullinated Autoantigens in Rheumatoid Arthritis Synovial Fluid. Arthritis Rheumatol, 2015;67:3135–45.

[7] A. Cross, T. Barnes, R. C. Bucknall, S. W. Edwards, R. J. Moots. Neutrophil apoptosis in rheumatoid arthritis is regulated by local oxygen tensions within joints. Journal of leukocyte biology, 2006;80:521–8.

[8] H. L. Wright, B. Chikura, R. C. Bucknall, R. J. Moots, S. W. Edwards. Changes in expression of membrane TNF, NF-{kappa}B activation and neutrophil apoptosis during active and resolved inflammation. Ann Rheum Dis, 2011;70:537–43.

[9] H. L. Wright, M. Lyon, E. A. Chapman, R. J. Moots, S. W. Edwards. Rheumatoid Arthritis Synovial Fluid Neutrophils Drive Inflammation Through Production of Chemokines, Reactive Oxygen Species, and Neutrophil Extracellular Traps. Front Immunol, 2020;11:584116.

[10] A. Cross, S. W. Edwards, R. C. Bucknall, R. J. Moots. Secretion of oncostatin M by neutrophils in rheumatoid arthritis. Arthritis Rheum, 2004;50:1430–6.

[11] L. Glennon-Alty, A. P. Hackett, E. A. Chapman, H. L. Wright. Neutrophils and redox stress in the pathogenesis of autoimmune disease. Free radical biology & medicine, 2018;125.

[12] C. Carmona-Rivera, P. M. Carlucci, R. R. Goel, E. James, S. R. Brooks, C. Rims et al. Neutrophil extracellular traps mediate articular cartilage damage and enhance cartilage component immunogenicity in rheumatoid arthritis. JCI Insight, 2020;5.

[13] C. Carmona-Rivera, P. M. Carlucci, E. Moore, N. Lingampalli, H. Uchtenhagen, E. James et al. Synovial fibroblast-neutrophil interactions promote pathogenic adaptive immunity in rheumatoid arthritis. Sci Immunol, 2017;2.

[14] H. L. Wright, T. Cox, R. J. Moots, S. W. Edwards. Neutrophil biomarkers predict response to therapy with tumor necrosis factor inhibitors in rheumatoid arthritis. Journal of leukocyte biology, 2017;101:785–95.

[15] H. L. Wright, H. B. Thomas, R. J. Moots, S. W. Edwards. Interferon gene expression signature in rheumatoid arthritis neutrophils correlates with a good response to TNFi therapy. Rheumatology (Oxford, England), 2015;54:188–93.

[16] A. Cross, R. C. Bucknall, M. A. Cassatella, S. W. Edwards, R. J. Moots. Synovial fluid neutrophils transcribe and express class II major histocompatibility complex molecules in rheumatoid arthritis. Arthritis Rheum, 2003;48:2796–806.

[17] U. Karmakar, S. Vermeren. Crosstalk between B cells and neutrophils in rheumatoid arthritis. Immunology, 2021;164:689–700.

[18] D. Minns, K. J. Smith, G. Hardisty, A. G. Rossi, E. Gwyer Findlay. The Outcome of Neutrophil-T Cell Contact Differs Depending on Activation Status of Both Cell Types. Front Immunol, 2021;12:633486.

[19] A. Moffat, E. Gwyer Findlay. Evidence for antigen presentation by human neutrophils. Blood, 2024;143:2455–63.

[20] R. Khandpur, C. Carmona-Rivera, A. Vivekanandan-Giri, A. Gizinski, S. Yalavarthi, J. S. Knight et al. NETs are a source of citrullinated autoantigens and stimulate inflammatory responses in rheumatoid arthritis. Sci Transl Med, 2013;5:178ra40.

[21] E. Pieterse, N. Rother, C. Yanginlar, J. Gerretsen, S. Boeltz, L. E. Munoz et al. Cleaved N-terminal histone tails distinguish between NADPH oxidase (NOX)-dependent and NOX-independent pathways of neutrophil extracellular trap formation. Ann Rheum Dis, 2018;77:1790–8.

[22] E. A. Chapman, M. Lyon, D. Simpson, D. Mason, R. J. Beynon, R. J. Moots et al. Caught in a Trap? Proteomic analysis of neutrophil extracellular traps in rheumatoid arthritis and systemic lupus erythematosus. Frontiers in Immunology, 2019;doi: 10.3389/fimmu.2019.00423.

[23] A. L. Cross, H. L. Wright, J. Choi, S. W. Edwards, N. Ruiz-Opazo, V. L. M. Herrera. Circulating Neutrophil Extracellular Trap (NET)-forming neutrophils in rheumatoid arthritis exacerbation are majority dual endothelin-1/signal peptide receptor (DEspR)+ subtype. Clinical and experimental immunology, 2024.

[24] J. Fransen, P. L. van Riel. The Disease Activity Score and the EULAR response criteria. Clinical and experimental rheumatology, 2005;23:S93–9.

[25] L. Glennon-Alty, R. J. Moots, S. W. Edwards, H. L. Wright. Type I interferon regulates cytokine-delayed neutrophil apoptosis, reactive oxygen species production and chemokine expression. Clinical and experimental immunology, 2021;203:151–9.

[26] H. L. Wright, H. B. Thomas, R. J. Moots, S. W. Edwards. RNA-Seq Reveals Activation of Both Common and Cytokine-Specific Pathways following Neutrophil Priming. PloS one, 2013;8:e58598.

[27] D. Kim, B. Langmead, S. L. Salzberg. HISAT: a fast spliced aligner with low memory requirements. Nat Methods, 2015;12:357–60.

[28] Y. Liao, G. K. Smyth, W. Shi. The R package Rsubread is easier, faster, cheaper and better for alignment and quantification of RNA sequencing reads. Nucleic Acids Res, 2019;47:e47.

[29] R Core Team. R: A language and environment for statistical computing. R Foundation for Statistical Computing, Vienna, Austria, 2021.

[30] M. D. Robinson, D. J. McCarthy, G. K. Smyth. edgeR: a Bioconductor package for differential expression analysis of digital gene expression data. Bioinformatics, 2010;26:139–40.

[31] M. E. Ritchie, B. Phipson, D. Wu, Y. Hu, C. W. Law, W. Shi et al. limma powers differential expression analyses for RNA-sequencing and microarray studies. Nucleic Acids Res, 2015;43:e47.

[32] J. T. Leek, W. E. Johnson, H. S. Parker, E. J. Fertig, A. E. Jaffe, Y. Zhang et al. sva: Surrogate Variable Analysis. R package version 3520, 2024.

[33] A. H. Patil, M. K. Halushka. miRge3.0: a comprehensive microRNA and tRF sequencing analysis pipeline. NAR Genom Bioinform, 2021;3:lqab068.

[34] A. Kozomara, M. Birgaoanu, S. Griffiths-Jones. miRBase: from microRNA sequences to function. Nucleic Acids Res, 2019;47:D155–D62.

[35] B. Langmead, C. Trapnell, M. Pop, S. L. Salzberg. Ultrafast and memory-efficient alignment of short DNA sequences to the human genome. Genome Biol, 2009;10:R25.

[36] M. I. Love, W. Huber, S. Anders. Moderated estimation of fold change and dispersion for RNA-seq data with DESeq2. Genome Biol, 2014;15:550.

[37] T. Wu, E. Hu, S. Xu, M. Chen, P. Guo, Z. Dai et al. clusterProfiler 4.0: A universal enrichment tool for interpreting omics data. Innovation (Camb), 2021;2:100141.

[38] G. Yu, L. G. Wang, Y. Han, Q. Y. He. clusterProfiler: an R package for comparing biological themes among gene clusters. OMICS, 2012;16:284–7.

[39] D. Croft, G. O’Kelly, G. Wu, R. Haw, M. Gillespie, L. Matthews et al. Reactome: a database of reactions, pathways and biological processes. Nucleic Acids Res, 2011;39:D691–7.

[40] J. Zyla, M. Marczyk, T. Domaszewska, S. H. E. Kaufmann, J. Polanska, J. Weiner. Gene set enrichment for reproducible science: comparison of CERNO and eight other algorithms. Bioinformatics, 2019;35:5146–54.

[41] D. Chaussabel, C. Quinn, J. Shen, P. Patel, C. Glaser, N. Baldwin et al. A modular analysis framework for blood genomics studies: application to systemic lupus erythematosus. Immunity, 2008;29:150–64.

[42] A. A. Margolin, I. Nemenman, K. Basso, C. Wiggins, G. Stolovitzky, R. Dalla Favera et al. ARACNE: an algorithm for the reconstruction of gene regulatory networks in a mammalian cellular context. BMC Bioinformatics, 2006;7 Suppl 1:S7.

[43] P. Shannon, A. Markiel, O. Ozier, N. S. Baliga, J. T. Wang, D. Ramage et al. Cytoscape: a software environment for integrated models of biomolecular interaction networks. Genome Res, 2003;13:2498–504.

[44] J. H. Morris, L. Apeltsin, A. M. Newman, J. Baumbach, T. Wittkop, G. Su et al. clusterMaker: a multi-algorithm clustering plugin for Cytoscape. BMC Bioinformatics, 2011;12:436.

[45] S. Maere, K. Heymans, M. Kuiper. BiNGO: a Cytoscape plugin to assess overrepresentation of gene ontology categories in biological networks. Bioinformatics, 2005;21:3448–9.

[46] A. Kramer, J. Green, J. Pollard, Jr., S. Tugendreich. Causal analysis approaches in Ingenuity Pathway Analysis. Bioinformatics, 2014;30:523–30.

[47] S. Hanzelmann, R. Castelo, J. Guinney. GSVA: gene set variation analysis for microarray and RNA-seq data. BMC Bioinformatics, 2013;14:7.

[48] A. Subramanian, P. Tamayo, V. K. Mootha, S. Mukherjee, B. L. Ebert, M. A. Gillette et al. Gene set enrichment analysis: a knowledge-based approach for interpreting genome-wide expression profiles. Proc Natl Acad Sci U S A, 2005;102:15545–50.

[49] Y. Zuo, S. Yalavarthi, H. Shi, K. Gockman, M. Zuo, J. A. Madison et al. Neutrophil extracellular traps in COVID-19. JCI Insight, 2020;5.

[50] S. Saithong, W. Saisorn, P. Tovichayathamrong, G. Filbertine, P. Torvorapanit, H. L. Wright et al. Anti-Inflammatory Effects and Decreased Formation of Neutrophil Extracellular Traps by Enoxaparin in COVID-19 Patients. Int J Mol Sci, 2022;23.

[51] H. L. Wright, F. A. Makki, R. J. Moots, S. W. Edwards. Low-density granulocytes: functionally distinct, immature neutrophils in rheumatoid arthritis with altered properties and defective TNF signalling. Journal of leukocyte biology, 2017;101:599–611.

[52] R. Singaravelu, N. Ahmed, C. Quan, P. Srinivasan, C. J. Ablenas, D. G. Roy et al. A conserved miRNA-183 cluster regulates the innate antiviral response. J Biol Chem, 2019;294:19785–94.

[53] G. Wigerblad, Q. Cao, S. Brooks, F. Naz, M. Gadkari, K. Jiang et al. Single-Cell Analysis Reveals the Range of Transcriptional States of Circulating Human Neutrophils. J Immunol, 2022;209:772–82.

[54] R. Grieshaber-Bouyer, F. A. Radtke, P. Cunin, G. Stifano, A. Levescot, B. Vijaykumar et al. The neutrotime transcriptional signature defines a single continuum of neutrophils across biological compartments. Nat Commun, 2021;12:2856.

[55] T. L. W. Muskardin, T. B. Niewold. Type I interferon in rheumatic diseases. Nat Rev Rheumatol, 2018;14:214–28.

[56] T. S. Mitchell, R. J. Moots, H. L. Wright. Janus kinase inhibitors prevent migration of rheumatoid arthritis neutrophils towards interleukin-8, but do not inhibit priming of the respiratory burst or reactive oxygen species production. Clinical and experimental immunology, 2017;189:250–8.

[57] S. Chokesuwattanaskul, M. Fresneda Alarcon, S. Mangalakumaran, R. Grosman, A. L. Cross, E. A. Chapman et al. Metabolic Profiling of Rheumatoid Arthritis Neutrophils Reveals Altered Energy Metabolism That Is Not Affected by JAK Inhibition. Metabolites, 2022;12.

[58] G. M. de Freitas Almeida, D. B. Oliveira, L. M. Botelho, L. K. Silva, A. C. Guedes, F. P. Santos et al. Differential upregulation of human 2’5’OAS genes on systemic sclerosis: Detection of increased basal levels of OASL and OAS2 genes through a qPCR based assay. Autoimmunity, 2014;47:119–26.

[59] E. Croze. Differential gene expression and translational approaches to identify biomarkers of interferon beta activity in multiple sclerosis. J Interferon Cytokine Res, 2010;30:743–9.

[60] Y. Sanayama, K. Ikeda, Y. Saito, S. Kagami, M. Yamagata, S. Furuta et al. Prediction of therapeutic responses to tocilizumab in patients with rheumatoid arthritis: biomarkers identified by analysis of gene expression in peripheral blood mononuclear cells using genome-wide DNA microarray. Arthritis Rheumatol, 2014;66:1421–31.

[61] L. Q. Qiu, P. Cresswell, K. C. Chin. Viperin is required for optimal Th2 responses and T-cell receptor-mediated activation of NF-kappaB and AP-1. Blood, 2009;113:3520–9.

[62] A. S. Gizzi, T. L. Grove, J. J. Arnold, J. Jose, R. K. Jangra, S. J. Garforth et al. A naturally occurring antiviral ribonucleotide encoded by the human genome. Nature, 2018;558:610–4.

[63] H. Kambara, F. Niazi, L. Kostadinova, D. K. Moonka, C. T. Siegel, A. B. Post et al. Negative regulation of the interferon response by an interferon-induced long non-coding RNA. Nucleic Acids Res, 2014;42:10668–80.

[64] R. El-Diwany, M. Soliman, S. Sugawara, F. Breitwieser, A. Skaist, C. Coggiano et al. CMPK2 and BCL-G are associated with type 1 interferon-induced HIV restriction in humans. Sci Adv, 2018;4:eaat0843.

[65] P. Arumugam, M. Chauhan, T. Rajeev, R. Chakraborty, K. Bisht, M. Madan et al. The mitochondrial gene-CMPK2 functions as a rheostat for macrophage homeostasis. Front Immunol, 2022;13:935710.

[66] J. Rodriguez-Carrio, M. Alperi-Lopez, P. Lopez, F. J. Ballina-Garcia, A. Suarez. Heterogeneity of the Type I Interferon Signature in Rheumatoid Arthritis: A Potential Limitation for Its Use As a Clinical Biomarker. Front Immunol, 2017;8:2007.

[67] P. K. Yadalam, T. Sivasankari, S. Rengaraj, M. H. Mugri, M. Sayed, S. S. Khan et al. Gene Interaction Network Analysis Reveals IFI44L as a Drug Target in Rheumatoid Arthritis and Periodontitis. Molecules, 2022;27.

[68] D. Zheng, T. Liwinski, E. Elinav. Inflammasome activation and regulation: toward a better understanding of complex mechanisms. Cell Discov, 2020;6:36.

[69] R. Ma, T. P. Ortiz Serrano, J. Davis, A. D. Prigge, K. M. Ridge. The cGAS-STING pathway: The role of self-DNA sensing in inflammatory lung disease. FASEB J, 2020;34:13156–70.

[70] F. Apel, L. Andreeva, L. S. Knackstedt, R. Streeck, C. K. Frese, C. Goosmann et al. The cytosolic DNA sensor cGAS recognizes neutrophil extracellular traps. Sci Signal, 2021;14.

[71] B. Frade-Sosa, A. Ponce, E. Ruiz-Ortiz, N. De Moner, M. J. Gomara, A. B. Azuaga, et al. Neutrophilic Activity Biomarkers (Plasma Neutrophil Extracellular Traps and Calprotectin) in Established Patients with Rheumatoid Arthritis Receiving Biological or JAK Inhibitors: A Clinical and Ultrasonographic Study. Rheumatol Ther, 2024;11:501–21.

[72] S. Dambal, M. Shah, B. Mihelich, L. Nonn. The microRNA-183 cluster: the family that plays together stays together. Nucleic Acids Res, 2015;43:7173–88.

[73] S. Li, W. Meng, Z. Guo, M. Liu, Y. He, Y. Li et al. The miR-183 Cluster: Biogenesis, Functions, and Cell Communication via Exosomes in Cancer. Cells, 2023;12.

[74] L. Long, Y. Deng, F. Yao, D. Guan, Y. Feng, H. Jiang et al. Recruitment of phosphatase PP2A by RACK1 adaptor protein deactivates transcription factor IRF3 and limits type I interferon signaling. Immunity, 2014;40:515–29.

[75] V. Shanker, G. Trincucci, H. M. Heim, H. T. Duong. Protein phosphatase 2A impairs IFNalpha- induced antiviral activity against the hepatitis C virus through the inhibition of STAT1 tyrosine phosphorylation. J Viral Hepat, 2013;20:612–21.

[76] Z. Wang, R. Dai, S. A. Ahmed. MicroRNA-183/96/182 cluster in immunity and autoimmunity. Front Immunol, 2023;14:1134634.

[77] K. Ichiyama, A. Gonzalez-Martin, B. S. Kim, H. Y. Jin, W. Jin, W. Xu et al. The MicroRNA-183- 96-182 Cluster Promotes T Helper 17 Cell Pathogenicity by Negatively Regulating Transcription Factor Foxo1 Expression. Immunity, 2016;44:1284–98.

[78] X. Wang, G. Wang, X. Zhang, Y. Dou, Y. Dong, D. Liu et al. Inhibition of microRNA-182-5p contributes to attenuation of lupus nephritis via Foxo1 signaling. Exp Cell Res, 2018;373:91–8.

[79] K. Theilgaard-Monch, L. C. Jacobsen, R. Borup, T. Rasmussen, M. D. Bjerregaard, F. C. Nielsen et al. The transcriptional program of terminal granulocytic differentiation. Blood, 2005;105:1785–96.

[80] M. P. Boldin, K. D. Taganov, D. S. Rao, L. Yang, J. L. Zhao, M. Kalwani et al. miR-146a is a significant brake on autoimmunity, myeloproliferation, and cancer in mice. J Exp Med, 2011;208:1189–201.

[81] A. Hawez, A. Al-Haidari, R. Madhi, M. Rahman, H. Thorlacius. MiR-155 Regulates PAD4- Dependent Formation of Neutrophil Extracellular Traps. Front Immunol, 2019;10:2462.

[82] O. I. Aruoma, B. Halliwell. Action of hypochlorous acid on the antioxidant protective enzymes superoxide dismutase, catalase and glutathione peroxidase. Biochem J, 1987;248:973–6.

[83] J. M. Adrover, C. Del Fresno, G. Crainiciuc, M. I. Cuartero, M. Casanova-Acebes, L. A. Weiss et al. A Neutrophil Timer Coordinates Immune Defense and Vascular Protection. Immunity, 2019;50:390–402 e10.

