## Supplementary Data for "Complexity of the neutrophil transcriptome in early and severe rheumatoid arthritis. A role for microRNAs?"

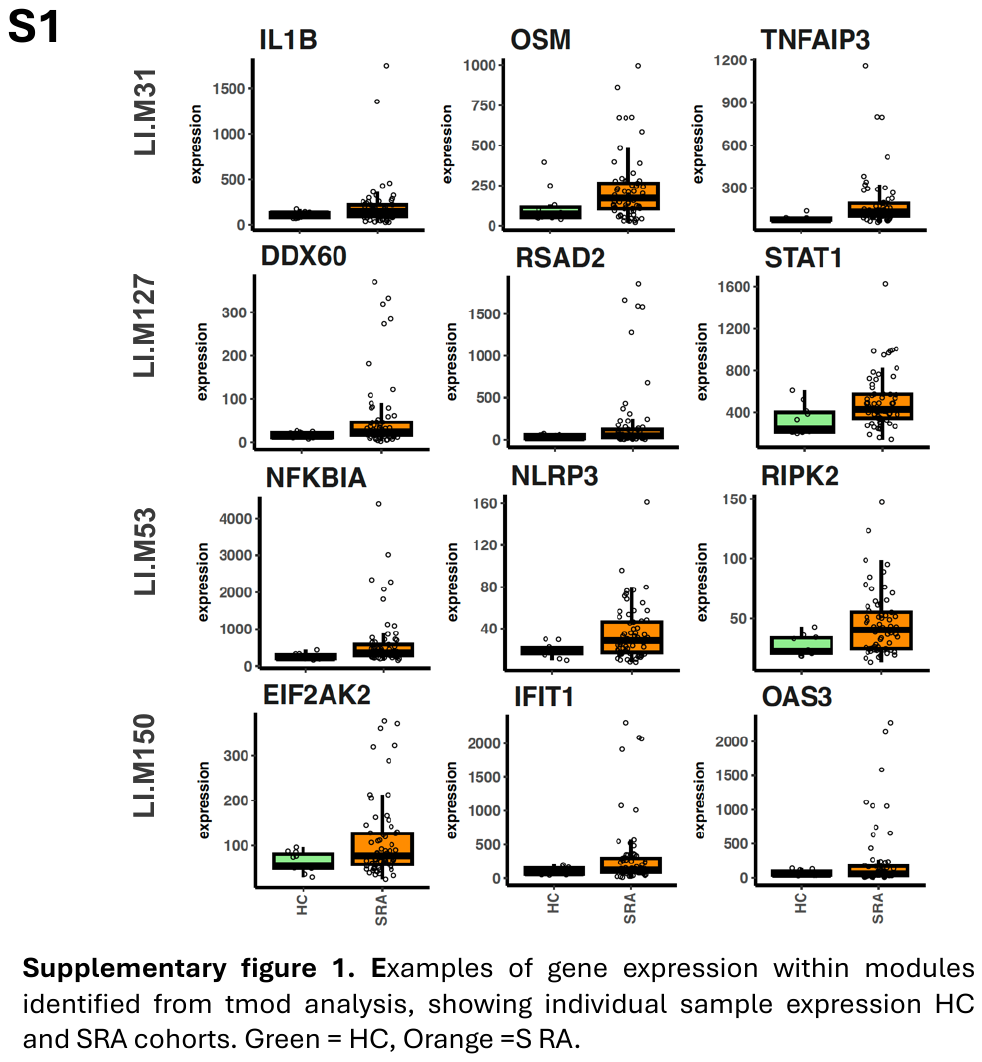

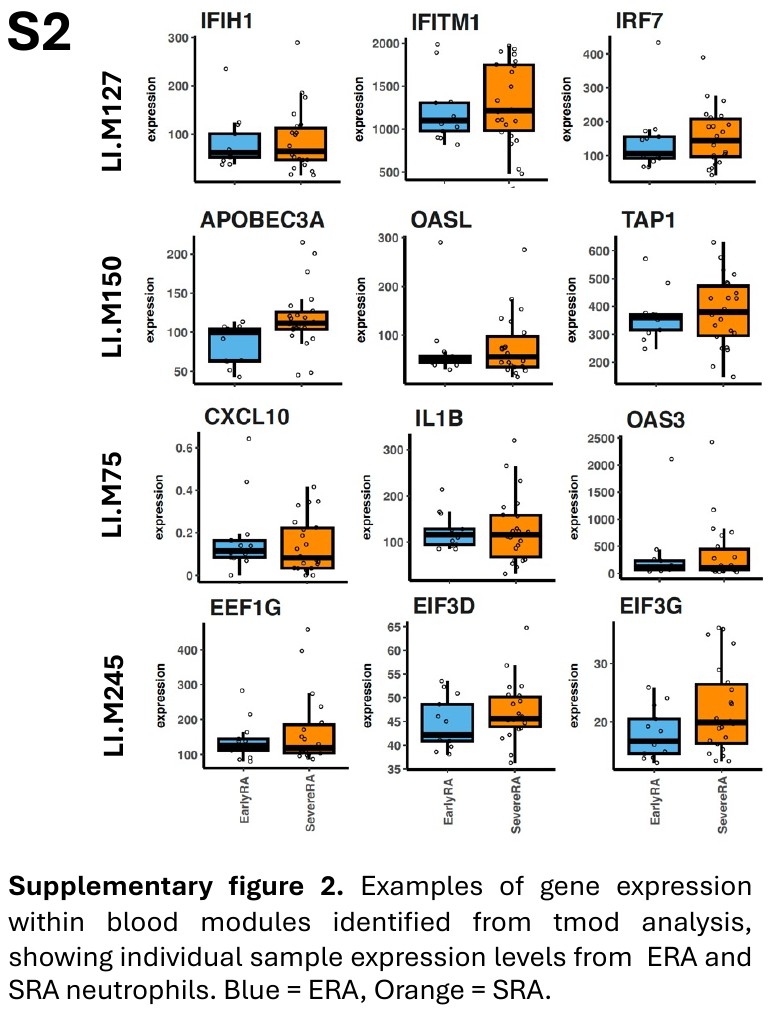

**Supplementary Table 1 – Upstream regulator analysis results for SRA vs HC mRNA.** Summary of predicted upstream cytokines and transcription regulators shown.

| **Upstream Regulator** | **Molecule Type** | **Predicted Activation State** | **Activation z-score** | **p-value of overlap** |
| --- | --- | --- | --- | --- |
| STAT1 | transcription regulator | Activated | 3.905 | 1.72E-27 |
| IFNG | cytokine | Activated | 6.17 | 5.46E-26 |
| IL1B | cytokine | Activated | 5.284 | 3.64E-23 |
| IFNL1 | cytokine | Activated | 4.381 | 1.15E-20 |
| NONO | transcription regulator | Activated | 4.507 | 1.90E-20 |
| ETV3 | transcription regulator | Inhibited | -3.5 | 2.84E-18 |
| IRF3 | transcription regulator | Activated | 4.093 | 2.00E-17 |
| TNF | cytokine | Activated | 4.911 | 9.61E-17 |
| IRF7 | transcription regulator | Activated | 4.693 | 2.86E-16 |
| SPI1 | transcription regulator | Activated | 2.182 | 3.19E-16 |
| IFNA2 | cytokine | Activated | 4.318 | 9.42E-15 |
| CSF3 | cytokine | Activated | 3.191 | 7.81E-14 |
| CREBBP | transcription regulator | Activated | 2.071 | 5.52E-13 |
| ETV6 | transcription regulator | Inhibited | -2.673 | 8.30E-13 |
| IFNB1 | cytokine | Activated | 3.454 | 9.37E-13 |
| IL1RN | cytokine | Inhibited | -2.192 | 4.58E-11 |
| PRL | cytokine | Activated | 2.793 | 4.88E-11 |
| IRF9 | transcription regulator | Activated | 3.148 | 1.14E-10 |
| IL6 | cytokine | Activated | 4.126 | 1.69E-10 |
| CITED2 | transcription regulator | Inhibited | -3.783 | 3.54E-10 |
| CD40LG | cytokine | Activated | 2.643 | 3.56E-10 |
| IRF1 | transcription regulator | Activated | 3.902 | 1.03E-09 |
| KLF6 | transcription regulator | Activated | 2.553 | 1.53E-09 |
| IL27 | cytokine | Activated | 3.921 | 2.41E-09 |
| IFNA1/IFNA13 | cytokine | Activated | 2.599 | 6.59E-09 |
| NKX2-3 | transcription regulator | Inhibited | -3.523 | 1.09E-08 |
| EPO | cytokine | Activated | 2.377 | 1.54E-08 |
| IL21 | cytokine | Activated | 3.85 | 2.76E-08 |
| RELA | transcription regulator | Activated | 2.351 | 2.76E-08 |
| NCOR1 | transcription regulator | Inhibited | -2.985 | 1.99E-07 |
| BHLHE40 | transcription regulator | Activated | 3.464 | 2.07E-07 |
| ZBTB10 | transcription regulator | Activated | 3.582 | 2.93E-07 |
| TRIM24 | transcription regulator | Inhibited | -3.132 | 4.62E-07 |
| ELK1 | transcription regulator | Activated | 2.358 | 1.42E-06 |
| IRF5 | transcription regulator | Activated | 2.746 | 3.18E-06 |
| PRDM1 | transcription regulator | Inhibited | -2.5 | 3.71E-06 |
| OSM | cytokine | Activated | 3.015 | 9.01E-06 |
| TWIST1 | transcription regulator | Activated | 2.557 | 9.01E-06 |
| IL5 | cytokine | Activated | 2.101 | 1.16E-05 |
| TAL1 | transcription regulator | Inhibited | -2.205 | 1.53E-05 |
| CREB1 | transcription regulator | Activated | 2.363 | 1.75E-05 |
| EGR1 | transcription regulator | Activated | 2.132 | 1.86E-05 |
| ARNT | transcription regulator | Activated | 2.156 | 1.91E-05 |
| FOXA2 | transcription regulator | Activated | 2.828 | 5.43E-05 |
| IL1A | cytokine | Activated | 2.765 | 5.80E-05 |
| PRDM16 | transcription regulator | Inhibited | -2.391 | 6.50E-05 |
| HIF1A | transcription regulator | Activated | 2.932 | 7.62E-05 |
| FOXL2 | transcription regulator | Activated | 2 | 1.86E-04 |
| SMARCB1 | transcription regulator | Activated | 2.531 | 2.11E-04 |
| IFNE | cytokine | Activated | 2.236 | 2.84E-04 |
| RB1 | transcription regulator | Inhibited | -2.017 | 3.65E-04 |
| HMGB1 | transcription regulator | Activated | 2.621 | 3.88E-04 |
| SP110 | transcription regulator | Inhibited | -2.121 | 7.45E-04 |
| SMAD3 | transcription regulator | Activated | 2.42 | 8.86E-04 |
| ETV7 | transcription regulator | Inhibited | -2 | 8.94E-04 |
| CEBPD | transcription regulator | Activated | 2.191 | 1.01E-03 |
| NFKB2 | transcription regulator | Activated | 2.2 | 2.35E-03 |
| DACH1 | transcription regulator | Inhibited | -2 | 2.50E-03 |
| IKZF3 | transcription regulator | Inhibited | -2.236 | 2.82E-03 |
| PPRC1 | transcription regulator | Activated | 2 | 3.49E-03 |
| CCL5 | cytokine | Activated | 2 | 4.47E-03 |
| VHL | transcription regulator | Inhibited | -2.177 | 4.70E-03 |
| EDN1 | cytokine | Activated | 2.28 | 8.74E-03 |
| IL7 | cytokine | Activated | 2.433 | 1.59E-02 |
| NUPR1 | transcription regulator | Activated | 2.516 | 2.62E-02 |
| YBX1 | transcription regulator | Activated | 2 | 4.27E-02 |

**Supplementary Table 2 – Upstream regulator analysis results for ERA vs HC mRNA.** Summary of predicted upstream cytokines and transcription regulators shown.

| **Upstream Regulator** | **Molecule Type** | **Predicted Activation State** | **Activation z-score** | **p-value of overlap** |
| --- | --- | --- | --- | --- |
| IL4 | cytokine | Inhibited | -2.505 | 6.13E-37 |
| ETV3 | transcription regulator | Inhibited | -5.385 | 4.80E-35 |
| IFNL1 | cytokine | Activated | 4.769 | 1.90E-33 |
| IFNG | cytokine | Activated | 5.288 | 6.92E-29 |
| STAT1 | transcription regulator | Activated | 3.5 | 1.12E-28 |
| NONO | transcription regulator | Activated | 5.86 | 1.46E-27 |
| ETV6 | transcription regulator | Inhibited | -4.811 | 2.03E-27 |
| IRF7 | transcription regulator | Activated | 5.396 | 5.81E-27 |
| IFNA2 | cytokine | Activated | 5.124 | 3.22E-23 |
| IRF3 | transcription regulator | Activated | 5.189 | 1.02E-20 |
| MYC | transcription regulator | Inhibited | -4.619 | 1.30E-20 |
| MLXIPL | transcription regulator | Inhibited | -5.099 | 7.79E-19 |
| TNF | cytokine | Activated | 2.128 | 3.47E-18 |
| PRL | cytokine | Activated | 4.359 | 1.14E-17 |
| IL27 | cytokine | Activated | 4.193 | 1.21E-17 |
| IFNB1 | cytokine | Activated | 4.312 | 1.46E-16 |
| IRF9 | transcription regulator | Activated | 3.417 | 3.14E-16 |
| IL1B | cytokine | Activated | 3.068 | 7.25E-15 |
| SPI1 | transcription regulator | Activated | 3.837 | 7.50E-15 |
| NKX2-3 | transcription regulator | Inhibited | -4.163 | 1.25E-14 |
| IRF1 | transcription regulator | Activated | 4.274 | 4.67E-14 |
| CITED2 | transcription regulator | Inhibited | -2.838 | 1.77E-13 |
| MYCN | transcription regulator | Inhibited | -2.835 | 5.03E-13 |
| STAT2 | transcription regulator | Activated | 2.312 | 1.29E-12 |
| IL1RN | cytokine | Inhibited | -2.163 | 3.69E-12 |
| FOXC1 | transcription regulator | Activated | 3.841 | 4.56E-12 |
| ZBTB10 | transcription regulator | Activated | 2.101 | 1.19E-11 |
| TRIM24 | transcription regulator | Inhibited | -4.043 | 1.68E-11 |
| IFNA1/IFNA13 | cytokine | Activated | 3.537 | 1.47E-10 |
| TWIST1 | transcription regulator | Activated | 2.071 | 6.49E-10 |
| IRF4 | transcription regulator | Inhibited | -2.1 | 1.10E-09 |
| NCOR1 | transcription regulator | Inhibited | -2.72 | 3.47E-09 |
| TNFSF10 | cytokine | Activated | 2.701 | 2.50E-08 |
| AIRE | transcription regulator | Inhibited | -2.121 | 2.50E-08 |
| CSF3 | cytokine | Activated | 2.161 | 4.27E-08 |
| MSC | transcription regulator | Activated | 2.111 | 6.09E-08 |
| PRDM16 | transcription regulator | Inhibited | -3.091 | 6.58E-08 |
| IFNL3 | cytokine | Activated | 2.063 | 8.03E-08 |
| IFNL4 | cytokine | Activated | 2.403 | 1.03E-07 |
| STAT5B | transcription regulator | Inhibited | -2.238 | 1.52E-07 |
| IRF5 | transcription regulator | Activated | 3.384 | 2.18E-07 |
| PRDM1 | transcription regulator | Inhibited | -2.659 | 2.27E-07 |
| CD40LG | cytokine | Inhibited | -2.255 | 2.62E-07 |
| PML | transcription regulator | Activated | 3.843 | 1.73E-06 |
| TNFSF9 | cytokine | Inhibited | -2.414 | 3.84E-06 |
| GATA3 | transcription regulator | Inhibited | -2.811 | 4.51E-06 |
| EP300 | transcription regulator | Inhibited | -2 | 5.13E-06 |
| NFAT5 | transcription regulator | Inhibited | -2.049 | 5.19E-06 |
| KLF7 | transcription regulator | Activated | 2.231 | 8.57E-06 |
| IKZF3 | transcription regulator | Inhibited | -2.813 | 1.33E-05 |
| MIF | cytokine | Inhibited | -2.208 | 1.43E-05 |
| KMT2D | transcription regulator | Inhibited | -3.637 | 2.35E-05 |
| IFNA4 | cytokine | Activated | 2.574 | 7.27E-05 |
| GATA1 | transcription regulator | Inhibited | -2.446 | 3.83E-04 |
| CLOCK | transcription regulator | Activated | 2.137 | 8.98E-04 |
| IFNK | cytokine | Activated | 2 | 1.09E-03 |
| SMARCB1 | transcription regulator | Activated | 3.162 | 1.75E-03 |
| FOXA2 | transcription regulator | Activated | 2.183 | 4.70E-03 |
| ZBED6 | transcription regulator | Activated | 2 | 4.80E-03 |
| IL5 | cytokine | Inhibited | -2.374 | 6.67E-03 |
| SP1 | transcription regulator | Activated | 2.781 | 9.01E-03 |
| IFNE | cytokine | Activated | 2 | 1.11E-02 |
| BRCA1 | transcription regulator | Activated | 2.158 | 1.40E-02 |
| SOX11 | transcription regulator | Inhibited | -2.63 | 2.70E-02 |

**Supplementary Table 3 – ARACNE2 gene expression network modules.** Gene ontology over-representation analysis was performed using BINGO (GO) and canonical pathway enrichment was performed using IPA.

| **Module** | **Method used** | **Adjusted**  **p-value** |
| --- | --- | --- |
| **M1. Metabolism & Transcription**  **(880 nodes, 19,281 edges)** |  |  |
| Regulation of gene expression | GO | 0.00018665 |
| Nucleobase, nucleoside, nucleotide and nucleic acid metabolic process | GO | 0.00018665 |
| Cellular metabolic process | GO | 0.0027225 |
| Valine degradation I | IPA | 0.00295121 |
| EIF2 signaling | IPA | 0.00512861 |
| mRNA processing | GO | 0.014741 |
| RNA transport | GO | 0.017427 |
| Cellular response to stress | GO | 0.024697 |
| Establishment of protein localization | GO | 0.034844 |
| Protein modification by small protein conjugation or removal | GO | 0.039013 |
| **M2. Integrin & Cytokine Receptors**  **(786 nodes, 25,308 edges)** |  |  |
| Intracellular signal transduction | GO | 7.0754E-08 |
| Production of NO and ROS in Macrophages | IPA | 1.122E-07 |
| NFAT in regulation of the immune response | IPA | 3.7154E-06 |
| Integrin signaling | IPA | 4.0738E-06 |
| IL-8 Signaling | IPA | 4.4668E-06 |
| IL-6 Signaling | IPA | 5.2481E-06 |
| Signaling by Rho family GTPases | IPA | 9.7724E-06 |
| Post-translational protein modification | GO | 1.1237E-05 |
| HMGB1 signaling | IPA | 1.3183E-05 |
| NF-kappaB signaling | IPA | 3.9811E-05 |
| **M3. Kinase Signalling**  **(394 nodes, 1,855 edges)** |  |  |
| Cellular macromolecule metabolic process | GO | 2.1E-15 |
| Protein modification process | GO | 4.73E-09 |
| AMPK signaling | IPA | 8.3176E-07 |
| Establishment of protein localization | GO | 1.18E-06 |
| Vesicle-mediated transport | GO | 1.94E-06 |
| Cdc42 signaling | IPA | 6.3096E-06 |
| NF-kappaB signaling | IPA | 0.00026915 |
| CD40 signaling | IPA | 0.00085114 |
| 3-phosphoinositide biosynthesis | IPA | 0.00218776 |
| SAPK/JNK signaling | IPA | 0.00186209 |
| **M4. Interferon & Toll-like Receptors**  **(89 nodes, 295 edges)** |  |  |
| Interferon signaling | IPA | 3.9811E-23 |
| Response to virus | GO | 3.0548E-15 |
| Activation of IRF by cytosolic pattern recognition receptors | IPA | 5.0119E-14 |
| Immune response | GO | 4.3147E-07 |
| Retinoic acid mediated apoptosis signaling | IPA | 3.6308E-06 |
| Role of RIG1-like receptors in antiviral innate immunity | IPA | 2.5704E-05 |
| Death receptor signaling | IPA | 0.00044668 |
| Innate immune response | GO | 0.0011464 |
| Regulation of defense response | GO | 0.0080641 |
| Regulation of defense response to virus | GO | 0.090327 |
| **M5. Gene Expression**  **(211 nodes, 623 edges)** |  |  |
| EIF2 signaling | IPA | 1.2589E-75 |
| Regulation of eIF4 and p70S6K signaling | IPA | 3.1623E-21 |
| Phospholipase C signaling | IPA | 8.3176E-07 |
| Apoptosis signaling | IPA | 0.01348963 |
| Translational elongation | GO | 8.0121E-89 |
| Gene expression | GO | 1.2008E-39 |
| Cellular macromolecule biosynthetic process | GO | 1.8894E-38 |
| Cellular protein metabolic process | GO | 4.1766E-30 |
| Cellular metabolic process | GO | 2.8799E-19 |
| Leukocyte activation | GO | 9.2644E-05 |
| **M6. Metabolism**  **(189 nodes, 2,194 edges)** |  |  |
| 4-hydroxyproline degradation I | IPA | 0.01348963 |
| Proline biosynthesis I | IPA | 0.02630268 |
| Proline biosynthesis II (from arginine) | IPA | 0.03981072 |
| Arginine degradation VI (Arginase 2 pathway) | IPA | 0.03981072 |
| Fucose metabolic process | GO | 0.040412 |
| Carbohydrate metabolic process | GO | 0.20522 |
| Aminoglycan metabolic process | GO | 0.21765 |
| Hexose metabolic process | GO | 0.21765 |
| Monosaccharide catabolic process | GO | 0.21765 |
| Glycosaminoglycan metabolic process | GO | 0.36956 |

**Supplementary Table 4 – Upstream regulator analysis results for SRA vs HC mRNA.** Summary of predicted upstream microRNAs regulating SRA gene expression.

| **Upstream Regulator** | **Molecule Type** | **Predicted Activation State** | **Activation z-score** | **p-value of overlap** |
| --- | --- | --- | --- | --- |
| mir-183 | microRNA | Activated | 2.263 | 4.64E-09 |
| mir-96 | microRNA | Activated | 2.985 | 8.23E-09 |
| miR-6087 (and other miRNAs w/seed GAGGCGG) | microRNA | Activated | 2.333 | 9.55E-03 |
| mir-21 | microRNA | Inhibited | -2.484 | 1.05E-02 |

**Supplementary Table 5 – Upstream regulator analysis results for SRA mRNA:miRNA target filter analysis.** Summary of predicted upstream microRNAs and transcription regulators.

| **Upstream Regulator** | **Molecule Type** | **Predicted Activation State** | **Activation z-score** | **p-value of overlap** |
| --- | --- | --- | --- | --- |
| ECSIT | transcription regulator | Inhibited | -2.176 | 9.59E-11 |
| ETV3 | transcription regulator | Inhibited | -2.53 | 2.04E-10 |
| NONO | transcription regulator | Activated | 3.578 | 1.22E-09 |
| IRF3 | transcription regulator | Activated | 2.885 | 1.69E-09 |
| STAT1 | transcription regulator | Activated | 2.082 | 6.25E-09 |
| TWIST1 | transcription regulator | Activated | 2.706 | 1.82E-08 |
| mir-183 | microRNA | Activated | 2.8 | 5.61E-08 |
| IRF7 | transcription regulator | Activated | 3.223 | 9.93E-08 |
| IRF1 | transcription regulator | Activated | 2.047 | 1.27E-07 |
| mir-96 | microRNA | Activated | 2.63 | 1.87E-07 |
| ETV6 | transcription regulator | Inhibited | -2.781 | 3.60E-07 |
| miR-182-5p (and other miRNAs w/seed UUGGCAA) | mature microRNA | Inhibited | -2.63 | 2.45E-06 |
| TRIM24 | transcription regulator | Inhibited | -2.804 | 1.18E-05 |
| STAT3 | transcription regulator | Inhibited | -2.341 | 1.30E-05 |
| KLF6 | transcription regulator | Inhibited | -2.789 | 6.06E-05 |
| FOXC1 | transcription regulator | Activated | 2.828 | 1.86E-04 |
| KLF2 | transcription regulator | Activated | 2.199 | 2.10E-04 |
| NFKB1 | transcription regulator | Inhibited | -2.689 | 2.28E-04 |
| ZFP36 | transcription regulator | Activated | 2.211 | 5.31E-04 |
| NFAT5 | transcription regulator | Inhibited | -2.63 | 8.05E-04 |
| MSC | transcription regulator | Activated | 2 | 3.01E-03 |
| GPS2 | transcription regulator | Activated | 2 | 3.75E-03 |
| STAT4 | transcription regulator | Inhibited | -2.03 | 5.95E-03 |
| WBP2 | transcription regulator | Activated | 2.236 | 9.29E-03 |
| TCF7L2 | transcription regulator | Inhibited | -2.4 | 2.55E-02 |
| SMARCA4 | transcription regulator | Inhibited | -2.236 | 2.68E-02 |
| NFIC | transcription regulator | Inhibited | -2 | 3.86E-02 |
